# Cognition and metacognition in functional motor symptoms and functional seizures: A case-control study

**DOI:** 10.1101/2025.09.25.25336646

**Authors:** Susannah Pick, L.S. Merritt Millman, Esin Gun Gursoy, Yasmine Basamh, Jemima Uloyok-Job, Jessica Davies, Lauren Blunstone, Snigdha Bhuma, Jan Coebergh, Anthony S. David, Mark J. Edwards, Laura H. Goldstein, John Hodsoll, Mitul A. Mehta, Timothy R. Nicholson, Biba Stanton, Joel S. Winston, Matthew Hotopf, Trudie Chalder

## Abstract

**Background:** Cognitive symptoms are common in functional neurological disorder (FND), yet there is inconsistent evidence of impaired neurocognitive test performance in this population. We aimed to assess core cognitive functions and global metacognition in patients with functional seizures (FS) and functional motor symptoms (FMS), and to explore relationships between cognition, metacognition, and cognitive symptoms in these groups.

**Methods:** This single-centre case-control study recruited participants with FS (n=50) and FMS (n=50), in addition to age- and gender-matched healthy controls (HC, n=50), and clinical controls with depression and/or anxiety disorders (CC, n=50). The Cambridge Neuropsychological Test Automated Battery was used to assess response speed, working memory, executive functions, and social-emotional processing, with subjective confidence rated for each test. Validated measures quantified intellectual functioning, performance validity, and global cognitive symptoms.

**Findings:** Impaired performance was demonstrated in both FND groups for sustained attention (p=0.03-<0.001) and set-shifting (p=0.01-0.001), withstanding correction for response speed, education, intellectual functioning, and psychotropic medication. Performance validity was comparable between groups. The FND groups reported reduced confidence for sustained attention performance (p<0.001) and elevated cognitive symptoms (p<0.001). Executive performance deficits correlated with reduced test-specific confidence in FS/FMS (p=0.02-<0.001). In FMS, confidence for sustained attention correlated negatively with cognitive symptoms (p=0.002). Cognitive symptoms were associated with psychological/physical symptom load, quality-of-life, and/or general functioning in FND and CC groups (p=0.04-<0.001).

**Interpretation:** Patients with FS and FMS displayed deficits in executive functioning performance, with intact metacognitive awareness, alongside significant cognitive complaints. These neurocognitive features were associated with poorer clinical status, warranting interventions targeting cognitive control and/or cognitive symptoms in everyday life.

**Funding:** This study was funded by a Medical Research Council Career Development Award granted to SP [MR/V032771/1]. This project also represents independent research part-funded by the NIHR Biomedical Research Centre at South London and Maudsley NHS Foundation Trust and King’s College London.

## Introduction

Cognitive symptoms, such as forgetfulness, distractibility, and disrupted goal-driven behaviours, are common in patients with functional neurological disorder (FND).^1–3^ As primary symptoms in functional cognitive disorder (FCD),^3^ or occurring alongside other FND presentations,^2,3^ cognitive complaints have been associated with greater psychological symptom load and compromised quality of life.^2,4–6^ Despite this, empirical evidence of cognitive deficits remains inconclusive. Intact, impaired, or superior neurocognitive test performance has been reported in FND samples compared to neurological, psychiatric, and/or healthy controls, across several relevant domains (e.g., attentional control, executive function, social-emotional processing).^2,3,5,7,8^

Beyond core cognitive functions, emerging findings indicate potential differences in metacognition in FND.^3,9^ Metacognition refers to awareness and control of one’s own cognitive processes, and adaptive use of this knowledge in goal-driven behaviours.^10^ Metacognitive deficits are associated transdiagnostically with mental health symptoms and maladaptive behaviour, and may be important treatment targets across neuropsychiatric disorders.^11^ ‘Global metacognition’ involves broad self-evaluations of cognitive performance in a given task/domain, whereas ‘local metacognition’ relates to momentary assessments of specific decisions or actions within a task/activity.^9,11^ In FND, altered global metacognition has been indicated by reduced concordance between neurocognitive test performance and post-diction self-evaluations,^2,3,5,9^ and by the presence of elevated subjective cognitive symptoms in the absence of observable performance deficits.^2,3,7,9^ However, few high-quality experimental studies have examined local metacognition in FND, with inconsistent results.^9^

These disparate findings may be due to methodological issues, such as suboptimal consideration of confounding variables (e.g., education, medication, performance validity), the wide range and variable quality of measures employed, and differing comparison groups.^2,5,8,9^ Few studies have compared cognitive and metacognitive performance profiles in specific FND subgroups using the same measures, which could highlight shared and distinct cognitive/metacognitive features, with clinical and mechanistic relevance.

Within a larger project examining aetiological factors and mechanisms in functional seizures (FS) and functional motor symptoms (FMS),^12^ this study aimed to directly compare cognitive performance profiles and global metacognition in patients with a primary diagnosis of FS or FMS (not FCD). We sought to compare these groups to healthy controls without physical or mental health disorders (HC), and clinical controls (CC) with major depression and/or anxiety disorders. The objectives were to examine overall between-group differences (FS/FMS/CC/HC) and FND subgroup-specific (FS/FMS) outcome profiles in 1) executive functioning, working memory, and social-emotional processing performance, 2) test-specific post-diction confidence evaluations, and 3) global cognitive complaints. We also sought to explore within-group relationships between cognitive/metacognitive outcomes and clinical characteristics (FS/FMS/CC).

Compared to HC and CC, we predicted that the FS and/or FMS groups would display:

- Elevated global cognitive symptoms on a validated scale^2^.
- Altered performance on tests of:

a. executive functioning (i.e., diminished sustained attention, set-shifting, response inhibition),^5,7,8^
b. working memory (poorer performance),^5,7,8^
c. social-emotional processing (enhanced emotional bias/reduced emotion recognition),^13^
- Reduced (global) metacognitive confidence regarding their test performance.^9^

## Methods

### Study design and participants

This single-centre, case-control study was based at King’s College London (KCL), conducted within a larger research programme.^12^ Ethical approval was granted by the North-West Greater Manchester South National Health Service (NHS) Research Ethics Committee, United Kingdom (UK, Reference: IRAS 322652). Recruitment and data collection proceeded from 11/2023 to 04/2025.

Participants with FMS (n=50) or FS (n=50) were recruited from neuropsychiatry/neurology services at King’s College Hospital, South London and Maudsley, and St George’s University Hospital NHS Foundation Trusts (London, UK). Most of the FS (82%) and FMS (96%) groups self-referred to the study following adverts shared by clinicians and/or through FND Hope UK. When self-referring to the study, participants in the FND groups were asked to provide documentary evidence of an FND diagnosis from a specialist clinician. Healthy (n=50) and clinical (n=50) controls were recruited using advertisements on community webpages and participant registers. Control participants were selected to frequency-match groups for age and gender (1:1 cases/controls). An a-priori power calculation indicated that a between-group (FS/FMS/CC/HC) ANOVA/ANCOVA, used to test the primary hypotheses, would require a sample of 180 to achieve 80% power for detection of a small effect (f=0.1, p<0.05).

Inclusion criteria required that participants were aged 18-65 years, fluent in English, and had normal/corrected eyesight. FND samples were required to have a confirmed DSM-5^1^ diagnosis with FS *or* FMS as their most prominent symptom. Clinical controls met criteria for a diagnosis of major depression and/or an anxiety disorder, confirmed with a structured clinical interview (‘Materials and Measures’). We excluded candidates with active severe psychiatric disorder (i.e., severe alcohol/substance use disorder, psychosis) and neurological disease (e.g., neurodegenerative disorder, epilepsy). Other exclusion criteria were being unable to perform tasks due to significant physical/cognitive impairment, current participation in an interventional trial, lifetime diagnosis of FND (HC/CC), or the presence of concurrent FS and FMS within the previous 12 months (FND). Individuals with a primary diagnosis of FCD were ineligible.

## Materials and measures

### Clinical measures (interview, self-report questionnaires)

A medical history interview was conducted at baseline (SP/LSMM/SB), including modules from the *Quick Structured Clinical Interview for DSM-5 Disorders*^14^ (Psychosis, Alcohol/Substance Use Disorder, Affective Disorders, Anxiety Disorders). Clinical self-report questionnaires (Supplementary Table 1) included validated measures of depression, generalised anxiety, psychological/somatoform dissociation, physical symptoms, health-related quality-of-life, and general functioning.^12^ A study-specific scale was developed to quantify FND symptom severity/impact (Supplementary Table 2).

The *Cognitive Failures Questionnaire* (*CFQ*)^15^ assessed subjective cognitive symptoms over the preceding six months, such as everyday memory and attention lapses, and disrupted action sequences. Higher scores signify poorer subjective cognitive functioning. Satisfactory psychometric properties have been demonstrated for this measure.^15–17^

### Medical Symptom Validity Test (MSVT)^18^

The *MSVT* is a computerised test designed to examine neuropsychological test performance validity, with sound psychometric properties.^19,20^ Performance validity indicators are *Immediate* and *Delayed Recall* scores, and immediate-delayed *Consistency* scores (test failure indicated by scores ≤85%). This test was included to assess task engagement/negative response biases across groups.^21^

### Wechsler Abbreviated Scale of Intelligence – 2nd edition (WASI-II)^22^

The *WASI-II* two-subtest version was administered to test non-verbal and verbal intellectual abilities (*Matrix Reasoning/Vocabulary*) and to provide an estimated *Full-Scale Intellectual Quotient Score (WASI-II FSIQ-2).* This version has strong inter-rater reliability (.95-.99), internal consistency (0.94), and test-retest stability (.94).^22^

### Cambridge Neuropsychological Test Automated Battery (CANTAB) Connect^23^

The *CANTAB Connect* platform is a modified version of the original battery, administered using a touchscreen device. Selected subtests probed motor/cognitive response times, attention, executive functioning, social-emotional cognition, and memory (Table 1, Supplementary Table 3). The original battery has strong psychometric properties in clinical and non-clinical samples,^23,24^ although psychometric validation data for the *Connect* version is not currently available.

**Table 1.** CANTAB Connect subtests.

| **Subtest** | **Domain** | **Description** |
| --- | --- | --- |
| **Motor Screening (2 min)** | Sensorimotor speed | Trials consist of a coloured cross presented onscreen in an unpredictable spatial location. The location of the cross varies between trials. Participants are asked to manually tap the cross as fast as possible.<br>**Outcome variables:** Total Correct/Incorrect; Motor Mean Latency |
| **Reaction Time (3 min)** | Motor and cognitive response speed | A circle is presented at the bottom of the screen, with five circles at the top. One of the top circles lights up on each trial. Participants must press on the bottom circle until the top circle lights up, then manually tap the illuminated upper circle as quickly as possible.<br>**Outcome variables:** Total Error Score; Movement Time; Reaction Time |
| **Rapid Visual Information Processing (~7 min)** | Sustained attention | A series of digits is presented individually onscreen at a rate of 100 digits/minute. Participants are asked to identify target sequences of digits (e.g., 6-2-9) by manually tapping a button onscreen as fast as possible.<br>**Outcome variables:** Ability score; Total Misses; Probability of Hit; False Alarm; Response Latency |
| **Spatial Span (5 min)** | Visuospatial working memory | Sequences of squares are shown changing colour individually, with an unpredictable spatial pattern. On each trial, participants are asked to tap the squares in the sequence that they changed colour. The Forward version requires the sequence to be repeated in the same order. The Reverse version requires the sequence to be repeated backwards. The number of squares increases during the task from 2-9, increasing the difficulty level.<br>**Outcome variables:** Forward/Reverse errors; Forward/Reverse Span Length |
| **Intra-Extra Dimensional Set Shift (7 min)** | Executive functioning: Attentional set-shifting & cognitive flexibility | Participants aim to identify an implicit rule based on an array of pink shapes and white lines. Participants manually select the stimulus that they think aligns with the current rule. Auditory feedback indicates 'Correct' or 'Incorrect' responses. After six correct responses, the rule changes and the participant attempts to identify the new rule. Rule changes can be within a dimension (intra-dimensional set shift), or they shift to a different dimension (extra-dimensional set shift). Difficulty increases across the trials.<br>**Outcome variables:** Total Trials/Stages; Total/Adjusted Errors; Extra-Dimensional Shift Errors; Pre-Extra-Dimensional Shift Errors; Completed Stage Trials/Errors; Response Latency |
| **Stop Signal Task (14 min)** | Executive functioning: Response inhibition | Trials consist of an arrow presented centrally at the top of the screen, pointing to the right or left, with a right and left button presented below on the respective side. Participants respond by manually tapping the right or left button, based on the direction of the arrow. On some trials, a tone is presented. Participants are asked to withhold their response when they hear the tone (response inhibition). The stop-signal delay is variable and adapts to the participant's performance.<br>**Outcome variables:** Missed Trials; Errors (Go/Stop trials); Stop Signal Reaction Time |
| **Emotional Bias Task (4 min)** | Social Cognition (Emotional processing) | Trials consist of a briefly presented morphed emotional face (150msec). The face varies in intensity between two emotions (e.g., happy-angry). Participants respond by selecting which of the two emotions they perceived (e.g., happiness or anger). Three versions of the task were administered (happy-anger, happy-disgust, happy-sadness).<br>**Outcome variables:** Reaction Time; Bias Point – Proportion of Happy selections. |
| **Emotion Recognition Test (6-10 min)** | Social cognition (Emotional processing) | Trials consist of a briefly presented emotional face (200msec), displaying anger, sadness, fear, disgust, surprise, or happiness. Participants respond by manually tapping the emotion they perceived from an array of six emotion labels.<br>**Outcome variables:** Reaction Times; False Alarms; Total/Unbiased Hits |

### Metacognitive confidence ratings

Post-diction confidence ratings were obtained immediately after every test (1=Very poor performance; 2=Poor performance; 3=Below average; 4=Average; 5=Above average; 6=Superior; 7=Very superior). Such global metacognitive assessments have been used widely in other clinical disorders.^11^

### Procedure

The recruitment and screening process are detailed in the study protocol^12^. After obtaining written consent, the medical history interview was conducted remotely. Eligible participants were sent a link to the online questionnaires, to be completed within 48 hours of the research visit. The cognitive battery was administered in a quiet testing room, between 10am-12pm for most participants. The session commenced with the *WASI-II*, followed by the *CANTAB* battery (fixed order, Table 1), then the *MSVT*. Participants were provided with standardised instructions for each test. Abstinence from caffeine/nicotine for >2 hours before the visit was requested of all participants. Those on psychotropic medications that might alter cognition were asked to abstain for 12-24 hours, provided abstinence was unlikely to cause adverse consequences (self-report/judgement of principal investigator/research group). Participants received a £50 shopping voucher on completion of the session.

### Data processing and statistical analyses

Data analyses were conducted in SPSS (v29, IBM, 2022) by SP and EGG independently, following a pre-registered analysis plan (https://osf.io/grh89/). Assumptions were checked for each test, and if violated, suitable corrections implemented (‘Results’). We defined outliers as scores of 2.5 standard deviations (+/-) from the group mean for each variable. Analyses were rerun with outliers winsorised in sensitivity analyses. Some participants failed to complete tests due to technicalities, symptoms, or declining (‘Results’). There were no within-test missing data points because each test required all trials/elements to be completed to yield a valid score.

Categorical variables were examined with Fisher’s exact/chi-squared tests. One-way Analysis of Variance (ANOVA) was used for between-group comparisons with most continuous variables. Mixed-model ANOVA was used for the *CANTAB Emotion Recognition Test*, with group as the between-groups factor (FMS/FS/CC/HC) and emotion (happiness, disgust, anger, fear, sadness, surprise) as the within-subjects factor. Significant main effects/interactions were examined with Bonferroni or Games-Howell post-hoc tests for ANOVA/Welch’s ANOVA. Cramer’s V, eta-squared, partial eta-squared, and omega-squared were calculated as effect sizes, as appropriate. Analysis of Covariance (ANCOVA) was applied in sensitivity analyses to examine the influence of potentially relevant confounding variables, such as performance validity, education, intellectual functioning, and psychotropic medication. Results that did not withstand sensitivity analyses were considered uninterpretable and not considered further.

Exploratory Spearman’s correlations were run between variables that differed significantly between groups. Benjamini-Hochberg corrections were applied within each set of tests to control the false discovery rate (5%). Results remaining significant after correction are reported.

## Results

Sociodemographic and clinical characteristics of the 200 participants are presented in Table 2 (Supplementary Tables 4-5), including common psychotropic medications and comorbid diagnoses. The groups were comparable in age, gender, and handedness. Significant overall group differences were found for self-reported education, psychotropic medication, comorbid physical/mental health diagnoses, anxiety, depression, somatoform and psychological dissociation, physical symptoms, health-related quality-of-life, and work/social functioning. These results largely reflected expected differences between HC and FS/FMS/CC groups (https://osf.io/preprints/psyarxiv/8vtpe_v1). The CC and FND groups were similar in rates of physical health comorbidities (p=0.25), and anxiety/depression severity (p=0.50-0.55), but they differed significantly in education (p=0.01) and psychotropic medication use (p<0.001).

**Table 2.**
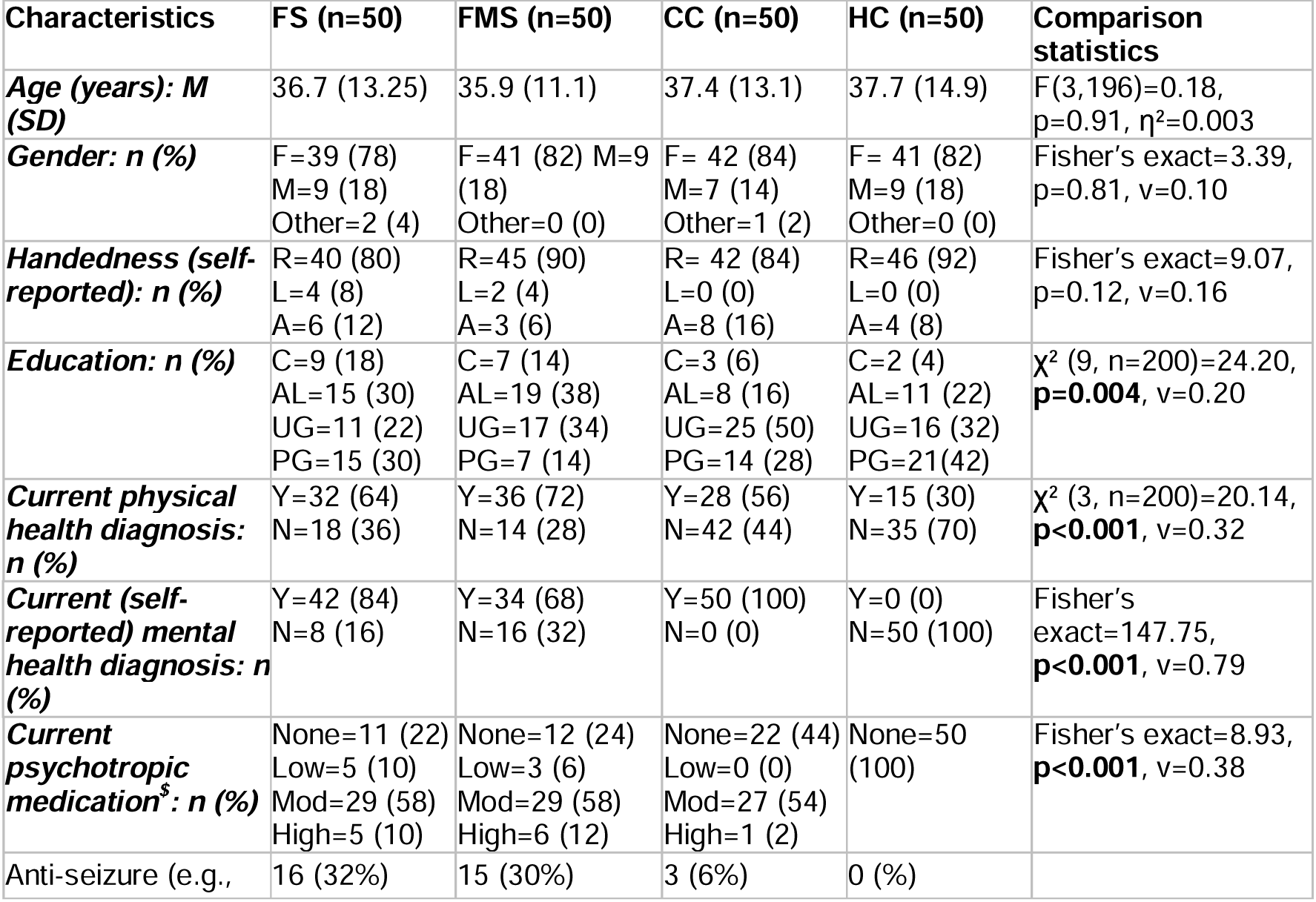
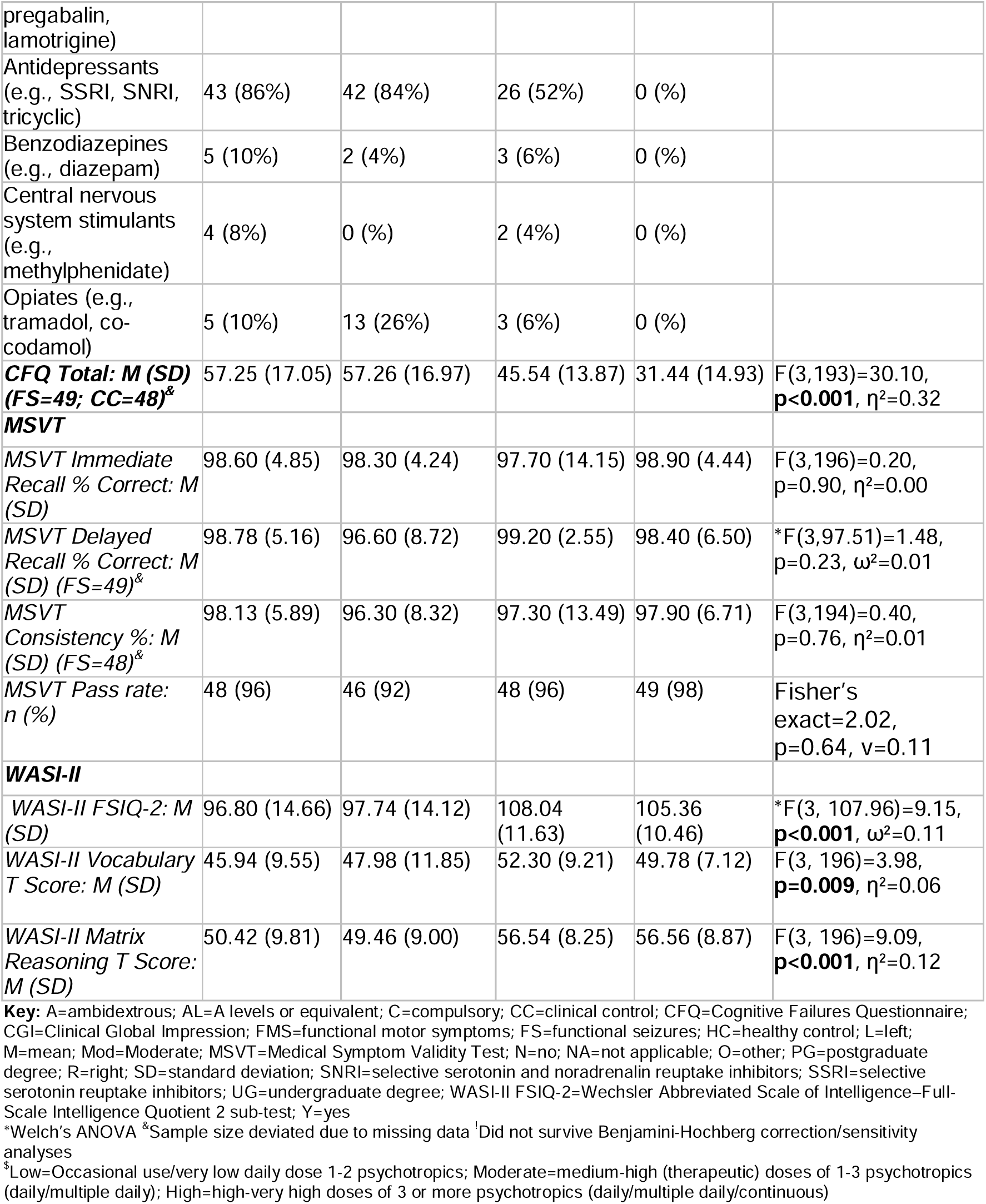
Sample characteristics, cognitive symptoms, performance validity, and intellectual functioning.

Figure 1 illustrates absolute effect sizes for the significant between-group differences in cognitive and metacognitive outcomes.

**Figure 1.**
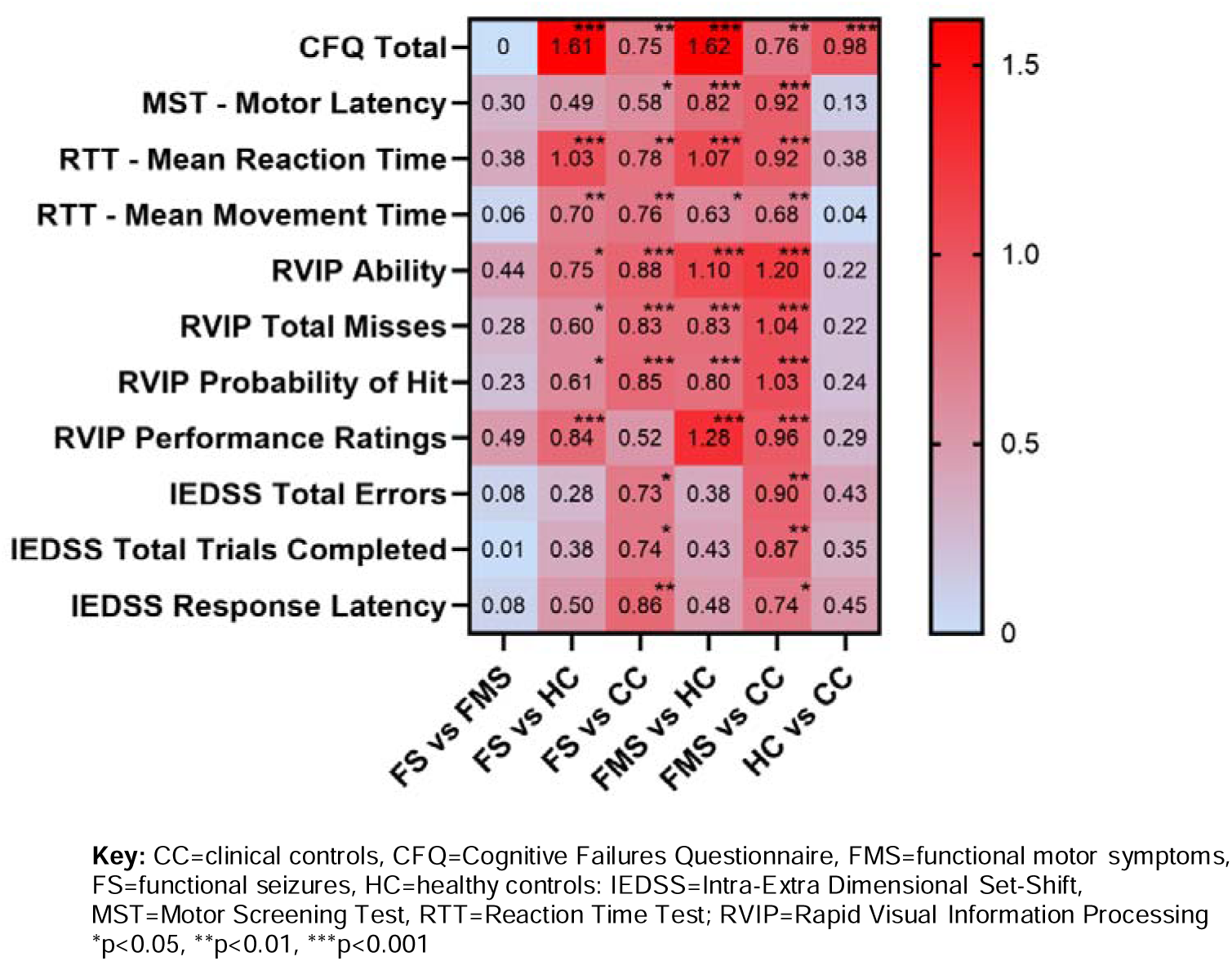
Cohen’s d (effect size) for cognitive symptoms (CFQ), Motor/Reaction Time (MST/RTT), Sustained Attention (RVIP) and Set-Shifting (IEDSS) tasks

Cognitive symptoms (*CFQ-Total*) were elevated in the FND and CC groups compared to HC, with scores also higher in the FND groups relative to CC (Table 2, Supplementary Table 6-7, p=0.002-<0.001). The *MSVT* performance validity *Pass* rate was comparable between groups (Table 2). On the *WASI-II* (Table 2, Supplementary Tables 8-9), mean *FSIQ-2* scores were in the average range for all groups. The FS group had lower scores than CC for *FSIQ-2* (p=0.005) and *Vocabulary* (p=0.006), whereas both FND groups displayed reduced performance on *Matrix Reasoning* compared to HC/CC (p=0.005-<0.001). Statistical values for CANTAB outcomes that varied consistently between groups are shown in Table 3 (Figures 1 and 2).

**Figure 2.**
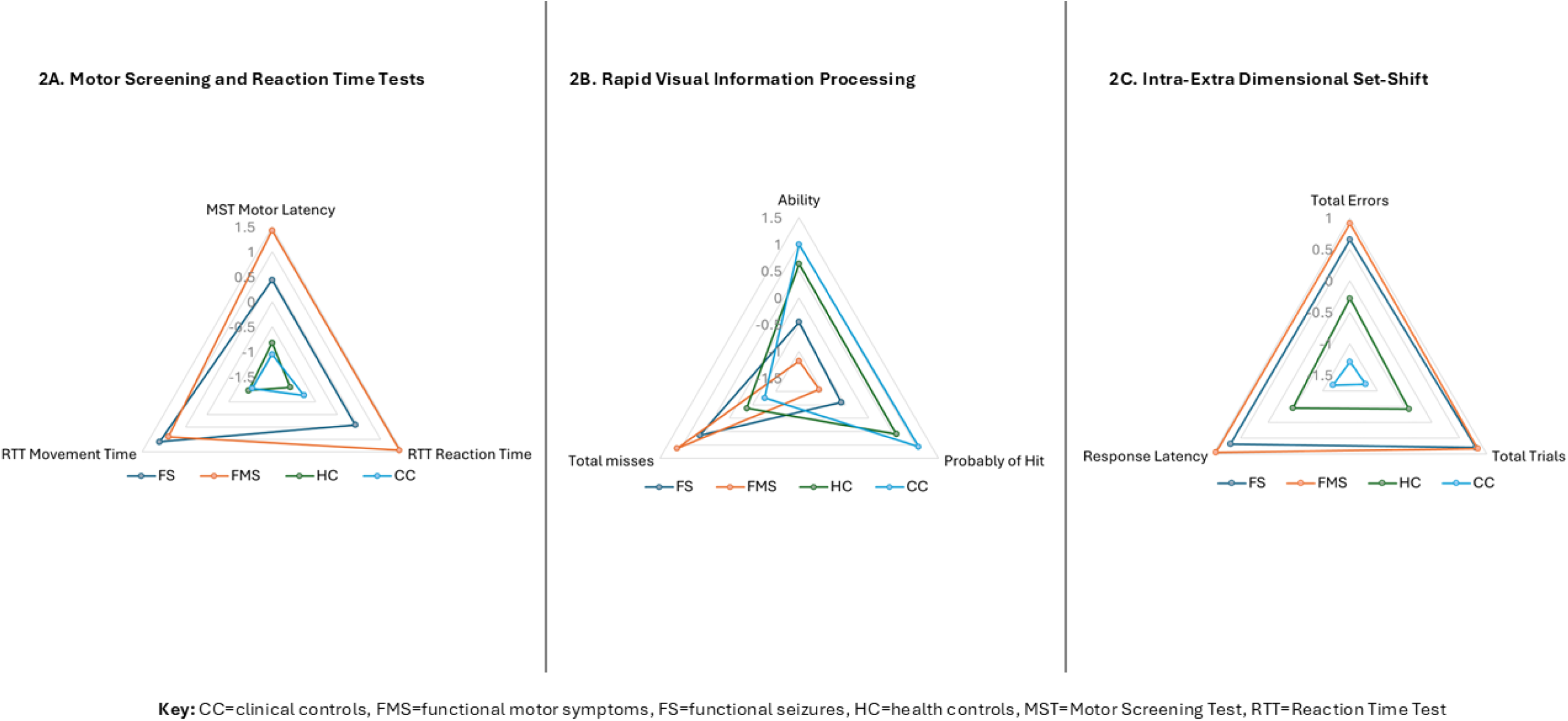
Radar plots - Motor Screening, Reaction Time, Rapid Visual Information Processing and Intra-Extra Dimensional Set-Shift subscale z-scores by group

**Table 3.** CANTAB results – Motor/reaction time, sustained attention, and set-shifting.

|  | FS | FMS | CC | HC | ANOVA statistics |
| --- | --- | --- | --- | --- | --- |
| **Motor Screening Test (MST)** | **n=50** | **n=50** | **n=50** | **n=50** |  |
| *MST - Mean Motor Latency (ms): M (SD)* | 913.55<br>(327.33) | 1016.04<br>(351.87) | 760.12<br>(178.29) | 783.94<br>(186.69) | $F(3,105.30)=8.88$,<br>**$p<0.001$**, $\omega^2=0.11$ |
| **Reaction Time Test (RTT)** | **n=50** | **n=50** | **n=50** | **n=50** |  |
| *RTT - Mean Reaction Time (ms): M (SD)* | 421.90<br>(84.15) | 466.16<br>(141.35) | 370.32<br>(40.71) | 356.43<br>(32.10) | $F(3,100.98)=16.81$,<br>**$p<0.001$**, $\omega^2=0.20$ |
| *RTT - Mean Movement Time (ms): M (SD)* | 297.05<br>(85.52) | 292.01<br>(86.38) | 243.95<br>(50.43) | 246.12<br>(57.23) | F (3,106.39)=7.96,<br>**p<0.001**, $\omega^2=0.10$ |
| **Rapid Visual Information Processing (RVIP)** | **n=47** | **n=50** | **n=49** | **n=49** |  |
| *RVIP Mdn Response Latency (ms): M (SD)* | 466.40<br>(92.40) | 543.07<br>(177.03) | 453.02<br>(80.91) | 457.07<br>(75.48) | *F(3,103.87)=3.73,<br>**p=0.01**, $\omega^2=0.08$ |
| *RVIP Ability: M (SD)* | 0.89<br>(0.04) | 0.87<br>(0.05) | 0.93<br>(0.05) | 0.92<br>(0.04) | F(3, 191)=13.40,<br>**p<0.001**, $\eta^2=0.17$ |
| *RVIP Total Misses: M (SD)* | 22.45<br>(8.55) | 25.04<br>(9.88) | 15.06<br>(9.28) | 17.10<br>(9.34) | F(3, 191)=12.22,<br>**p<0.001**, $\eta^2=0.16$ |
| *RVIP Probability of Hit: M (SD)* | 0.58<br>(0.16) | 0.54<br>(0.18) | 0.72<br>(0.17) | 0.68<br>(0.17) | F(3, 191)=12.22,<br>**p<0.001**, $\eta^2=0.16$ |
| *RVIP Probability of False Alarm: M (SD)* | 0.01<br>(0.03) | 0.02<br>(0.04) | 0.01<br>(0.01) | 0.01<br>(0.05) | F(3, 191)=0.82, p=0.49,<br>$\eta^2=0.01$ |
| **Intra-Extra Dimensional Set-Shift (IEDSS)** | **n=40** | **n=39** | **n=43** | **n=44** |  |
| *IEDSS Total Errors: M (SD)* | 18.20<br>(11.79) | 19.05<br>(10.65) | 11.84<br>(4.36) | 15.14<br>(10.02) | *F(3, 79.94)=7.99,<br>**p<0.001**, $\omega^2=0.07$ |
| *IEDSS Adjusted Errors: M (SD)* | 39.76<br>(49.35) | 32.80<br>(30.52) | 20.14<br>(22.77) | 27.28<br>(39.07) | *F(3,105.13)=3.18,<br>**p=0.03**, $\omega^2=0.02$ |
| *IEDSS Extra-Dimensional Shift Errors: M (SD)* | 5.81<br>(6.13) | 8.91<br>(8.93) | 7.02<br>(8.14) | 7.82<br>(9.89) | *F(3,100.54)=1.31,<br>p=0.28, $\omega^2=0.00$ |
| *IEDSS Pre-Extra-Dimensional Shift Errors: M (SD)* | 13.32<br>(11.03) | 12.90<br>(11.92) | 7.72<br>(6.90) | 10.42<br>(9.13) | *F(3,106.45)=4.29,<br>**p=0.007**, $\omega^2=0.04$ |
| *IEDSS Stages Completed: M (SD)* | 8.04<br>(2.08) | 8.36<br>(1.21) | 8.68<br>(0.84) | 8.46<br>(1.61) | *F(3,103.78)=1.78,<br>p=0.16, $\omega^2=0.01$ |
| *IEDSS Total Trials Completed: M (SD)* | 85.80<br>(23.98) | 86.05<br>(20.16) | 72.53<br>(9.54) | 77.75<br>(18.47) | *F(3, 82.04)=7.34,<br>**p<0.001**, $\omega^2=0.07$ |
| *IEDSS Completed Stage Trials: M (SD)* | 74.88<br>(29.61) | 77.82<br>(18.95) | 70.92<br>(11.75) | 71.90<br>(20.95) | *F(3,103.42)=1.69,<br>p=0.17, $\omega^2=0.00$ |
| *IEDSS Response Latency (ms): M (SD)* | 131119.23<br>(56928.56) | 136710.59<br>(81216.06) | 91660.98<br>(32183.23) | 107135.32<br>(37080.14) | *F(3, 84.39)=7.18,<br>**p<0.001**, $\omega^2=0.09$ |

**Key:** CC=clinical controls; FS=functional seizures; FMS=functional motor symptoms; HC=healthy controls; IEDSS=Intra-Extra Dimensional Set-Shift; M=mean; ms=milliseconds; MST=Motor Screening Test; RTT=Reaction Time Test; RVIP=Rapid Visual Information Processing; SD=standard deviation `Welch's ANOVA

*Motor Screening Test (MST) Mean Motor Latency* was extended in FMS compared to both HC and CC, and in FS relative to CCs (p=0.02-<0.001, Supplementary Tables 10-11).

*Reaction Time Test (RTT*) *Mean Reaction Time* and *Mean Movement Time* were elevated in both FND groups compared to HC/CC (p=0.01-<0.001). As extended response speed in the FND groups could unduly impair performance, *MST* and *RTT* variables were entered as covariates in sensitivity analyses for all other CANTAB outcomes.

Sustained attention performance differed significantly between groups on *RVIP Ability, Total Misses, Probability of Hit*, and *Median Response Latency* (Supplementary Tables 12-13), although the group effect on *RVIP Median Response Latency* did not endure sensitivity analyses (*MSVT* fails, *RTT Mean Reaction Time)*. Both FND groups displayed inferior performance to HC and CCs on the other *RVIP* outcomes (p=0.03-<0.001).

Set-shifting task performance deviated significantly between-groups on *IEDSS Total Errors, Adjusted Errors, Pre-Extra-Dimensional Shift Errors, Total Trials Completed*, and *Response Latency* (Supplementary Tables 14-15). The FS and FMS groups had more *Total Errors*, *Total Trials Completed* and extended *Response Latency,* compared to CCs (p-values 0.01-0.001). The group effects on *Adjusted Error* and *Pre-Extra-Dimensional Shift Errors* dissipated in sensitivity analyses (*MSVT* fails/education/*MST/RTT*).

None of the significant group main effects and/or interactions on *Spatial Span, Stop Signal, Emotion Bias, or Emotion Recognition* tests remained stable in sensitivity analyses (*MSVT/*education/*FSIQ-2*/psychotropic medication/*MST/RTT*, Supplementary Tables 16-22).

Both FND groups expressed significantly lower post-diction confidence for sustained attention (*RVIP*) performance than HCs, and the FMS group also reported reduced confidence on this task compared to CCs (p-values<0.001; Supplementary Tables 23-25). The groups did not differ in confidence ratings for any other test.

Within-groups (FS/FMS/CC) correlations (Supplementary Table 26) showed that impaired performance on the sustained attention (*RVIP*) and set-shifting (*IEDSS*) tests did not correlate significantly with global cognitive symptoms (*CFQ Total)* in any group. However, performance on these tests did correlate with test-specific confidence in the FMS and/or FS samples (p=0.01-<0.001). In the FMS group, significant associations were found between performance outcomes on sustained attention (*RVIP*) and set-shifting tasks (*IEDSS,* p=0.048-0.002).

Cognitive symptoms (*CFQ-Total*) correlated positively with clinical features in the FND/CC samples, including anxiety (r_s_=0.34-0.38, p=0.02-0.008), depression (r_s_=0.45-0.48, p=0.001-<0.001), and psychological/somatoform dissociation (r_s_=0.29-0.63, p=0.04-<0.001). *CFQ-Total* scores were also related to physical symptoms in the FS/CC groups (r_s_=0.39-0.45, p=0.006-0.001), and to greater FNS severity (r_s_=0.34, p=0.03) and impact (r_s_=0.30, p=0.02) in the FMS group. *CFQ-Total* scores were also associated with work/social functioning in the FMS/CC groups (r_s_=0.39-0.50, p=0.005-<0.001), and quality-of-life in FND/CC (r_s_=-0.33-0.57, p=0.03-<0.001).

In FMS, confidence ratings for sustained attention (*RVIP*) correlated with clinical variables, including cognitive symptoms (*CFQ-Total*, r_s_=-0.43, p=0.002), psychological dissociation (r_s_=-0.36-0.59, p=0.04-<0.001), depression (r_s_=-0.39, p=0.005), anxiety (r_s_=-0.37, p=0.007), and role limitations due to physical factors (r_s_=-0.54, p<0.001).

## Discussion

We examined cognitive and metacognitive functioning in patients with FMS or FS, compared to healthy and clinical controls (HC/CC). As predicted, the FND groups reported greater cognitive symptom burden, combined with localised performance impairments on tests of sustained attention and set-shifting. In contrast to our hypotheses, there were no consistent group differences in response inhibition, working memory, or social-emotional processing. In the FMS and/or FS groups, impaired performance in sustained attention and/or set-shifting was associated with reduced confidence ratings in those domains, suggesting broadly intact metacognitive accuracy.

Elevated cognitive complaints in our FS and FMS samples replicated several studies,^2,3,25,26^ showing that cognitive symptoms are a significant challenge for individuals with FND, beyond those with a primary diagnosis of FCD. As found in other populations, cognitive symptoms were related to a range of clinical features in our FS/FMS samples, but they did not correlate with performance deficits.^2,4,5,27,28^ Clinical correlates of cognitive symptoms included depression, anxiety, dissociation, physical symptoms, quality-of-life, and work/social functioning, with some of these associations more prominent in our FMS sample. However, these cognitive difficulties were heightened in our FS/FMS samples compared to CCs, who were comparable in anxiety and depression severity. Together, these findings underscore the prominence and potential impact of cognitive complaints in FND and provide preliminary evidence that these symptoms may be particularly relevant in patients with FMS.

Our results indicate that cognitive symptoms in FND may not be due solely to the presence of elevated anxiety and/or depression. Future research should evaluate the extent to which cognitive symptoms in FND are related to observable deficits in real-world settings, in addition to ascertaining the direction of influence between cognitive symptoms and other clinical outcomes. Importantly, patients with FMS and FS may require targeted interventions to facilitate management of cognitive symptoms and their impact in everyday life.^29^

Lower intellectual functioning and extended response speed were observed in both FND groups, corresponding with some previous studies,^5,8^ although intellectual functioning was qualitatively matched between groups, with mean scores in the average range. A minority of participants failed the performance validity test (*MSVT*), although the proportion was not significantly different between groups, consistent with other studies.^21^ Nonetheless, some of our primary results failed to survive sensitivity analyses including one or more of these variables, reinforcing the importance of assessing their influence in neuropsychological studies in future. The lack of robust group differences on response inhibition, working memory, and social-emotional processing tests countered our predictions, adding to the inconsistent findings in these domains.^5,8,13^ More thorough assessment of constituent processes with multiple tests may be necessary to uncover disorder-specific deficits in these functions.

Performance deficits on sustained attention (*RVIP*) and set-shifting (*IEDSS*) tasks exhibited by our FS and FMS samples withstood sensitivity analyses, therefore representing compelling evidence for specific executive function difficulties. The findings relating to sustained attention align with prior accounts of attentional control impairments in FND samples.^5,7,8,26^ Our observation of weaker performance on the set-shifting task (*IEDSS*) in the FND groups differed to two previous studies that used the same task,^2,30^ although the pattern of deficits suggested overall rule-learning difficulties during the task, rather than set-shifting performance specifically. Effect sizes were large for sustained attention performance in this sample, whereas they were small-medium for set-shifting, demonstrating that differences in the former domain might be more clinically meaningful.

Impaired attentional control might represent a valuable neurocognitive marker in FS and FMS, characterised by diminished maintenance of attentional focus, together with difficulties in learning implicit rules and/or voluntarily switching from a particular cognitive set. However, given that test performance in these domains did not correlate meaningfully with clinical outcomes (e.g., FND, cognitive, psychological, and physical symptoms, quality-of-life, work/social functioning), the relevance of these differences to FND should be scrutinised in greater detail. Investigators may also examine executive function performance in FS/FMS relative to patients with FCD, to identify possible shared and distinctive profiles and mechanisms in these subgroups.

Participants with FS and FMS expressed markedly reduced confidence on the sustained attention task, mirroring the performance deficit displayed on this measure. Actual performance and confidence ratings on the set-shifting task (*IEDSS*) were correlated within both FND groups, but not in CCs, indicating that the FND groups were aware of their difficulties to some extent. These results imply intact global metacognition in these domains in FS/FMS, contrary to our hypothesis and some prior findings,^3,5,7,9^ although more work is needed to establish this with greater certainty.

Confidence for sustained attention test performance, but not actual performance on that test, correlated significantly with global cognitive symptoms in the FMS group, replicating our pilot findings.^2^ Lower confidence for sustained attention performance was also associated with other adverse clinical characteristics in the FMS group. Therefore, reduced confidence in attentional control might be relevant to increased subjective cognitive symptoms and/or other clinical outcomes in FMS, beyond the degree of actual performance deficits displayed.

Further investigations should clarify the direction of effects between these variables, whilst also seeking to replicate these findings with additional neurocognitive tests.

The major strength of this study was the direct comparison of two common FND subgroups using the same test battery, with a sample size providing adequate statistical power to detect small effects. Inclusion of HC and CC groups provided insights into cognitive features in FND that were distinct from the general population, whilst controlling for elevated psychological distress. We employed a range of measures to comprehensively assess cognitive performance, along with two measures of global metacognition (subjective symptoms, post-diction appraisals), in addition to a test of performance validity. Our groups were comparable on most relevant background characteristics (e.g., age, gender, handedness), and we accounted for any pertinent group differences within our analyses (e.g., education, response speed, psychotropic medications). Examining potential relationships between cognitive variables and other clinical features allowed us to explore the clinical significance of neurocognitive differences in our FS/FMS/CC samples.

Some limitations of this study should be considered. Whilst we controlled statistically for the influence of education, psychotropic medications, and other confounds, there may have been residual effects on our results. A minority of our FND/CC samples reported diagnoses of attention-deficit hyperactivity disorder, which may have affected attentional performance. The use of a retrospective self-report questionnaire to assess cognitive symptoms may have resulted in recall biases. The *CANTAB Connect* battery lacks psychometric validation, established clinical cut-off scores, and has limited ecological validity. Performance-controlled experimental assessment of local metacognitive accuracy would complement measures of global metacognition in future studies.^9^

In conclusion, our findings provide robust evidence of performance deficits in sustained attention and set-shifting in FS and FMS, with seemingly intact global metacognitive awareness of these impairments, accompanied by elevated cognitive complaints in daily life. Poorer subjective cognitive functioning is associated with greater physical/psychological symptom burden, decreased work/social functioning, and worse quality-of-life, in patients with FND and/or common psychological symptoms (anxiety/depression). Neuropsychological rehabilitation approaches might hold promise for some patients with FMS and FS, specifically targeting cognitive control and associated self-appraisals in everyday activities, and facilitating self-management of subjective cognitive symptoms and their impact.

## Data Availability

The anonymised dataset will be made available in response to reasonable requests, on a case-by-case basis. Completion of a Data Sharing Agreement will be required, facilitated by the Kings College London Data Management team.

## Acknowledgements

We are grateful to all participants and the FND Patient and Carer Advisory Panel who oversaw the study. With thanks to FND Hope UK, clinicians and administrative staff at King’s College Hospital, South London and Maudsley, and St George’s University Hospital NHS Foundation Trusts (including Dr Simon Harrison, Dr Albert Joseph, Dr Sotiris Posporelis, Dr Norman Poole, Dr Tim Segal), and the RADAR-MDD team, for contributions to recruitment and screening. We thank NIHR BioResource volunteers for their participation, and gratefully acknowledge NIHR BioResource centres, NHS Trusts and staff for their contribution. Thanks also to the research assistants who assisted in some testing sessions (Chrystel-Jazz Campbell, Akshaana Manoharan).

## Supplementary Materials

**Supplementary Table 1.**
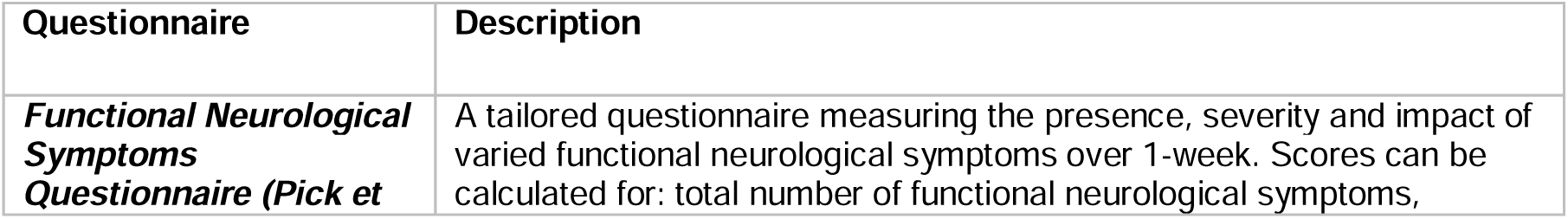
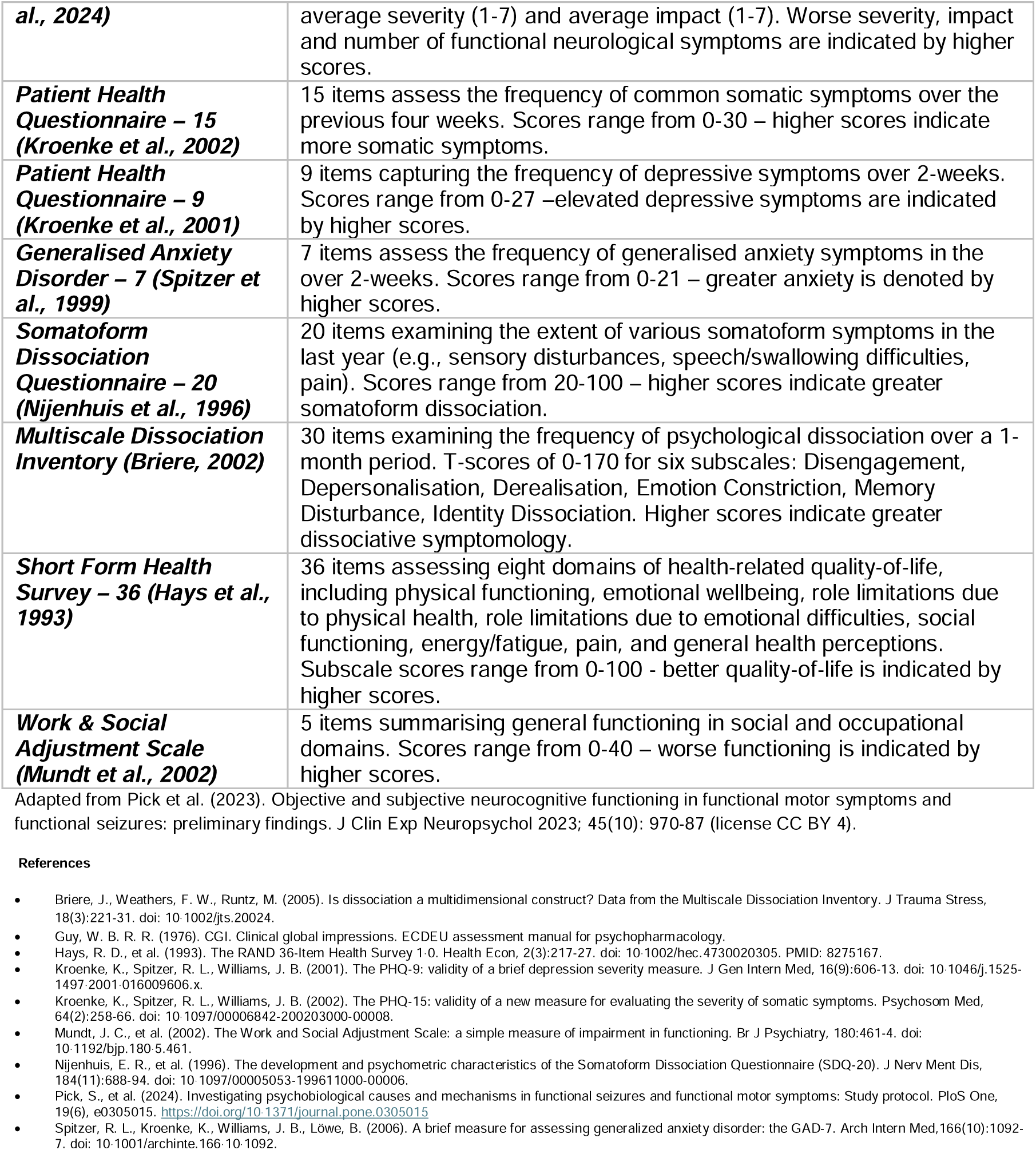
Clinical self-report questionnaire details.

**Supplementary Table 2.** Functional Neurological Symptoms Questionnaire (Pick et al., 2024) Please look at the symptoms in the table below and tell us whether you have experienced these functional neurological symptoms in the past week.LIf you mark yes to indicate that the symptom was present in the past week, please complete the additional columns to tell us how frequent the symptoms were, how severe (intense) they were, and how much impact they had on you. When rating the average severity of symptoms, please choose a number from 1 to 7, where 1=Symptom not present; 2=Minimal; 3=Mild; 4=Moderate; 5=Moderately severe; 6=Severe; 7=Very severe. When rating the impact of symptoms, please choose a number from 1 to 7, where 1=No impact at all; 2=Minimal impact; 3=Mild impact; 4=Moderate impact; 5=Moderately severe impact; 6=Severe impact; 7=Very severe impact.

| FND Symptom | Present?<br>(circle or bold) | Frequency<br>(circle or bold) | Average severity<br>(1-7) | Average impact<br>(1-7) |
| --- | --- | --- | --- | --- |
| Weakness | Yes / No | Constant / daily / weekly / less than weekly |  |  |
| Tremor | Yes / No | Constant / daily / weekly / less than weekly |  |  |
| Dystonia (muscle spasms / fixed postures) | Yes / No | Constant (1) / daily (2) / weekly (3) / less than weekly (4) |  |  |
| Walking / mobility difficulties | Yes / No | Constant / daily / weekly / less than weekly |  |  |
| Myoclonus (muscle jerks) | Yes / No | Constant / daily / weekly / less than weekly |  |  |
| Seizures* | Yes / No | Number of seizures in the last week: |  |  |
| Numbness (loss of feeling) | Yes / No | Constant / daily / weekly / less than weekly |  |  |
| Visual disturbances | Yes / No | Constant / daily / weekly / less than weekly |  |  |
| Sensitivity to light/sound | Yes / No | Constant / daily / weekly / less than weekly |  |  |
| Dizziness | Yes / No | Constant / daily / weekly / less than weekly |  |  |
| Speech / swallowing difficulties | Yes / No | Constant / daily / weekly / less than weekly |  |  |
| Cognitive difficulties (e.g., brain fog, memory lapses) | Yes / No | Constant / daily / weekly / less than weekly |  |  |
| Other FND symptoms | Details: | Constant / daily / weekly / less than weekly |  |  |

Please tell us which FND symptom(s) is most severe and has the most impact on you:
\*If you experience FND seizures, do you have warning symptoms? Yes / No
\*If you experience warning symptoms before an FND seizure, what is the earliest or most consistent symptom(s) that you experience?

**Supplementary Table 3.**
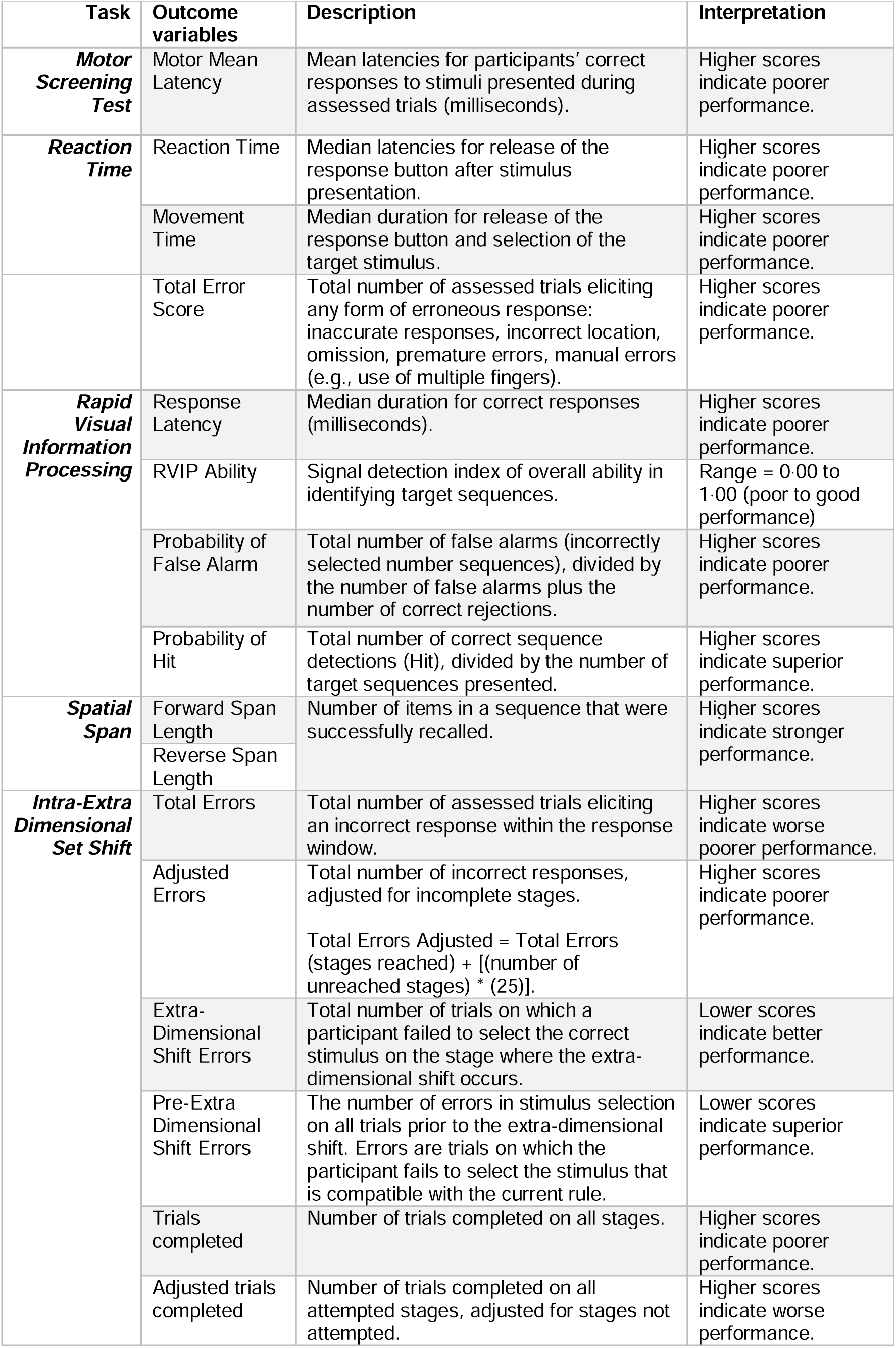
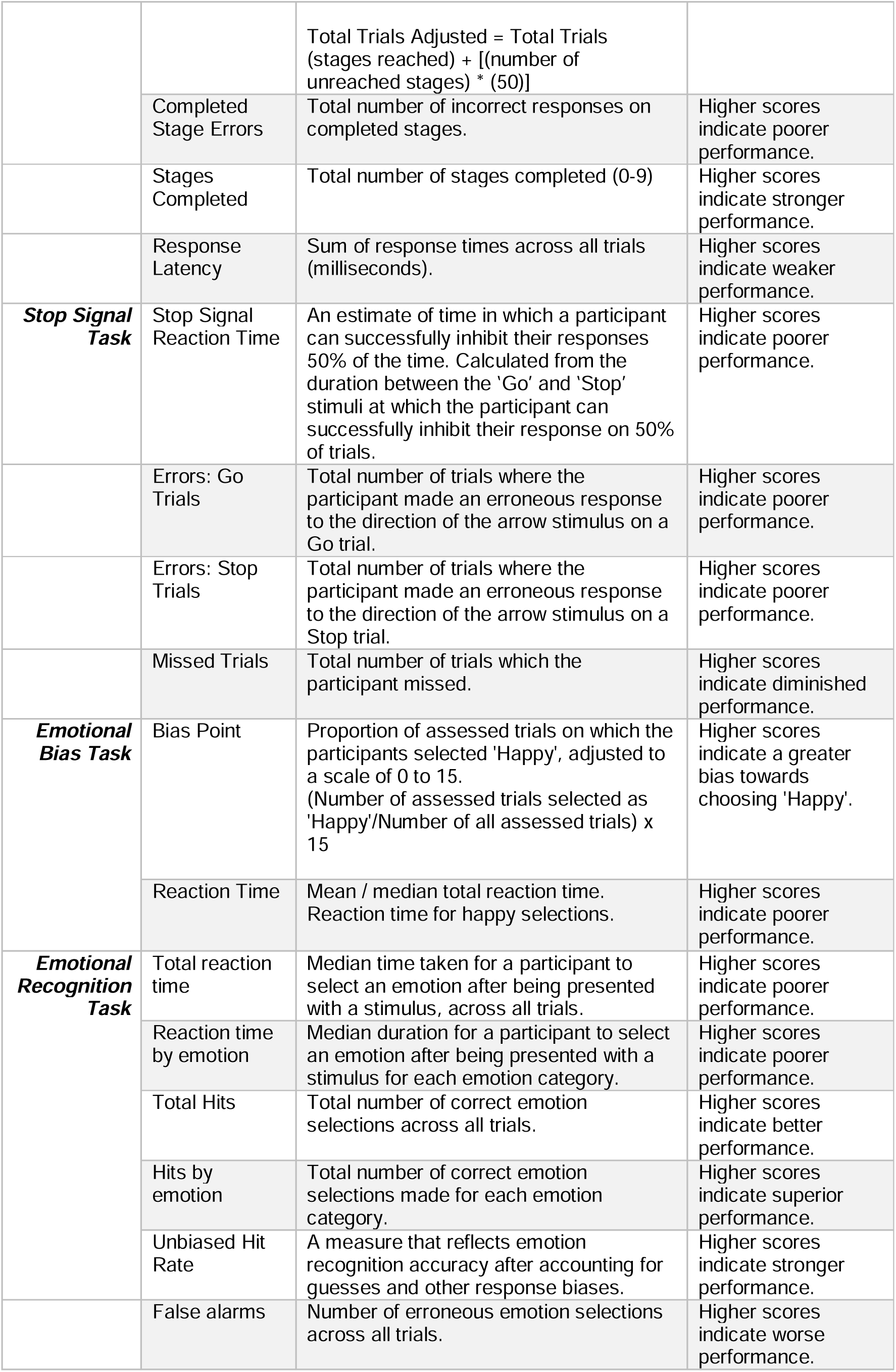
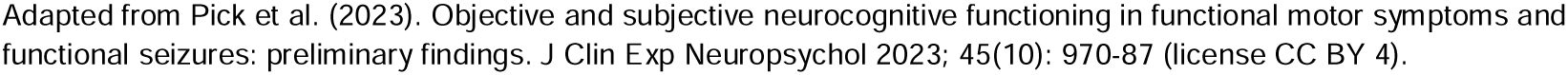
CANTAB Connect outcome variables.

**Supplementary Table 4.**
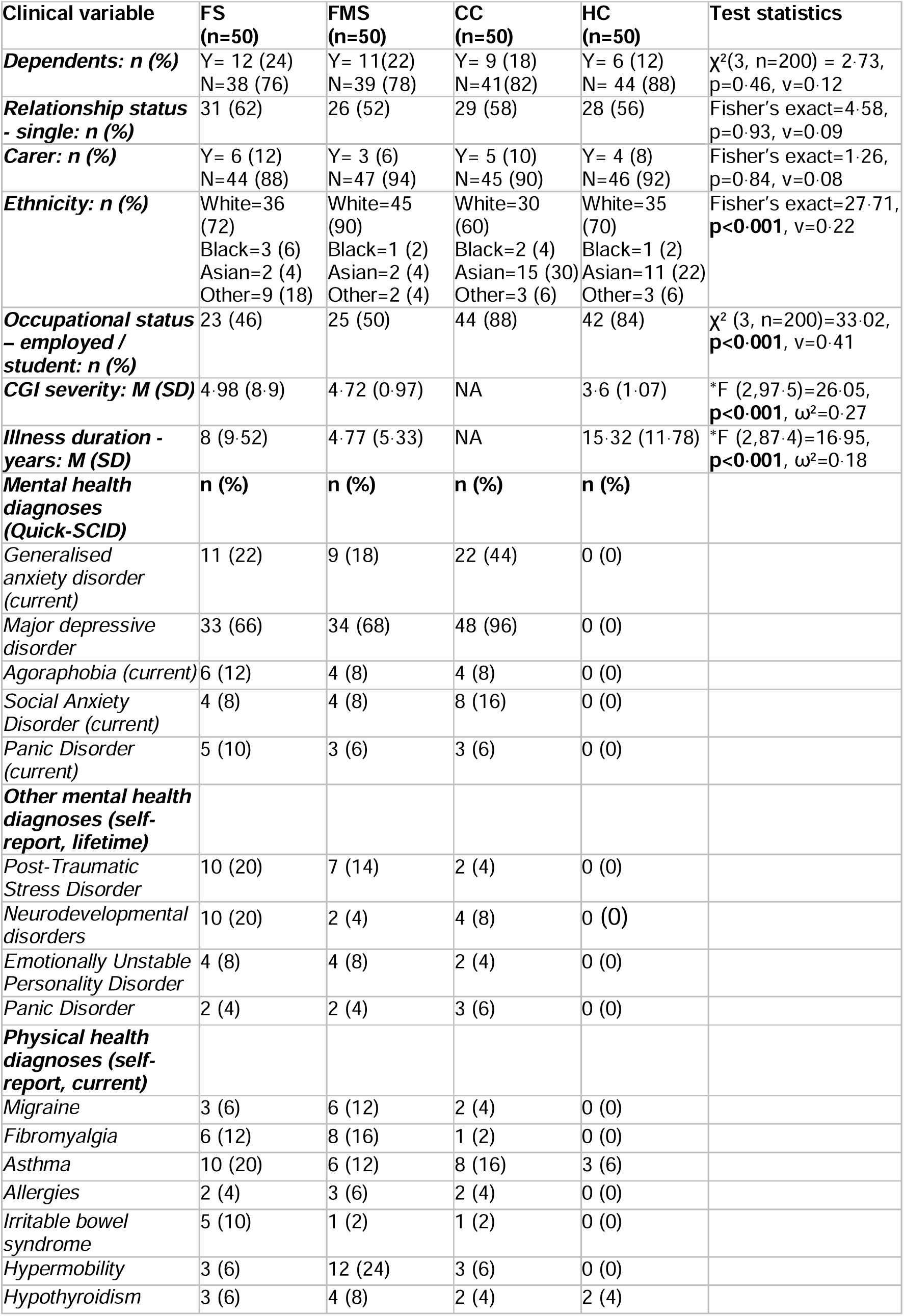
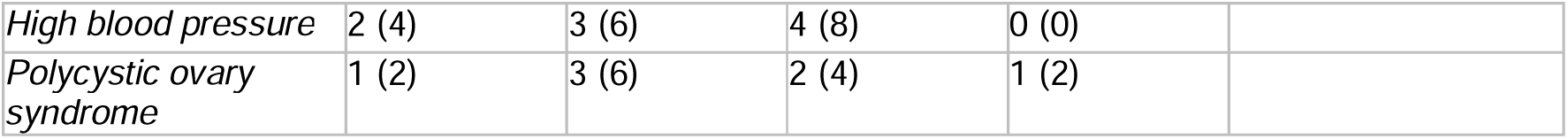
Sample characteristics: Sociodemographic and clinical variables.

**Supplementary Table 5.**
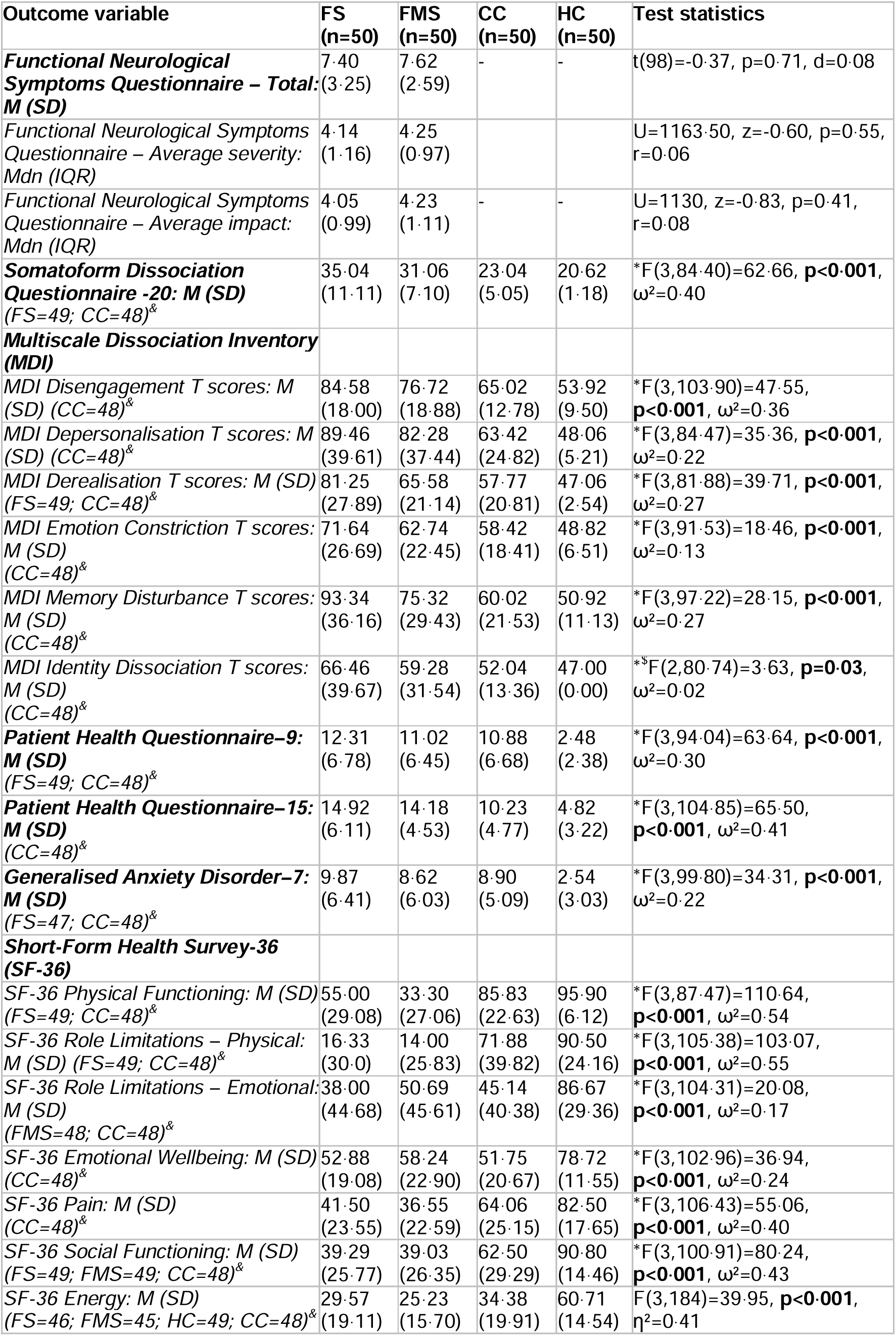
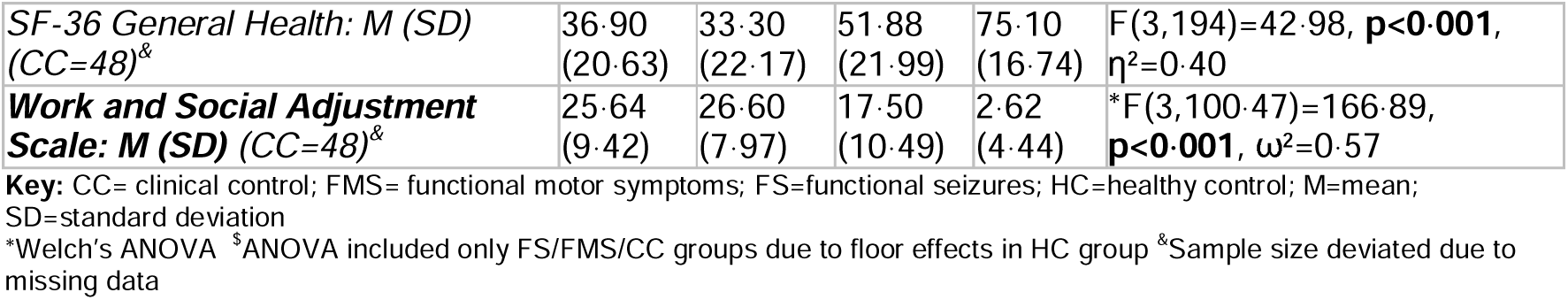
Clinical self-report measure statistics.

| Outcome variable | FS<br>(n=50) | FMS<br>(n=50) | CC<br>(n=50) | HC<br>(n=50) | Test statistics |
| --- | --- | --- | --- | --- | --- |
| **Functional Neurological Symptoms Questionnaire – Total: M (SD)** | 7.40<br>(3.25) | 7.62<br>(2.59) | - | - | t(98)=-0.37, p=0.71, d=0.08 |
| Functional Neurological Symptoms Questionnaire – Average severity: Mdn (IQR) | 4.14<br>(1.16) | 4.25<br>(0.97) |  |  | U=1163.50, z=-0.60, p=0.55, r=0.06 |
| Functional Neurological Symptoms Questionnaire – Average impact: Mdn (IQR) | 4.05<br>(0.99) | 4.23<br>(1.11) | - | - | U=1130, z=-0.83, p=0.41, r=0.08 |
| **Somatoform Dissociation Questionnaire -20: M (SD)**<br>(FS=49; CC=48)<sup>§</sup> | 35.04<br>(11.11) | 31.06<br>(7.10) | 23.04<br>(5.05) | 20.62<br>(1.18) | *F(3,84.40)=62.66, **p<0.001**, $\omega^2=0.40$ |
| **Multiscale Dissociation Inventory (MDI)** |  |  |  |  |  |
| MDI Disengagement T scores: M (SD) (CC=48)<sup>§</sup> | 84.58<br>(18.00) | 76.72<br>(18.88) | 65.02<br>(12.78) | 53.92<br>(9.50) | *F(3,103.90)=47.55, **p<0.001**, $\omega^2=0.36$ |
| MDI Depersonalisation T scores: M (SD) (CC=48)<sup>§</sup> | 89.46<br>(39.61) | 82.28<br>(37.44) | 63.42<br>(24.82) | 48.06<br>(5.21) | *F(3,84.47)=35.36, **p<0.001**, $\omega^2=0.22$ |
| MDI Derealisation T scores: M (SD) (FS=49; CC=48)<sup>§</sup> | 81.25<br>(27.89) | 65.58<br>(21.14) | 57.77<br>(20.81) | 47.06<br>(2.54) | *F(3,81.88)=39.71, **p<0.001**, $\omega^2=0.27$ |
| MDI Emotion Constriction T scores: M (SD) (CC=48)<sup>§</sup> | 71.64<br>(26.69) | 62.74<br>(22.45) | 58.42<br>(18.41) | 48.82<br>(6.51) | *F(3,91.53)=18.46, **p<0.001**, $\omega^2=0.13$ |
| MDI Memory Disturbance T scores: M (SD) (CC=48)<sup>§</sup> | 93.34<br>(36.16) | 75.32<br>(29.43) | 60.02<br>(21.53) | 50.92<br>(11.13) | *F(3,97.22)=28.15, **p<0.001**, $\omega^2=0.27$ |
| MDI Identity Dissociation T scores: M (SD) (CC=48)<sup>§</sup> | 66.46<br>(39.67) | 59.28<br>(31.54) | 52.04<br>(13.36) | 47.00<br>(0.00) | *F(2,80.74)=3.63, **p=0.03**, $\omega^2=0.02$ |
| **Patient Health Questionnaire–9: M (SD)**<br>(FS=49; CC=48)<sup>§</sup> | 12.31<br>(6.78) | 11.02<br>(6.45) | 10.88<br>(6.68) | 2.48<br>(2.38) | *F(3,94.04)=63.64, **p<0.001**, $\omega^2=0.30$ |
| **Patient Health Questionnaire–15: M (SD)**<br>(CC=48)<sup>§</sup> | 14.92<br>(6.11) | 14.18<br>(4.53) | 10.23<br>(4.77) | 4.82<br>(3.22) | *F(3,104.85)=65.50, **p<0.001**, $\omega^2=0.41$ |
| **Generalised Anxiety Disorder–7: M (SD)**<br>(FS=47; CC=48)<sup>§</sup> | 9.87<br>(6.41) | 8.62<br>(6.03) | 8.90<br>(5.09) | 2.54<br>(3.03) | *F(3,99.80)=34.31, **p<0.001**, $\omega^2=0.22$ |
| **Short-Form Health Survey-36 (SF-36)** |  |  |  |  |  |
| SF-36 Physical Functioning: M (SD) (FS=49; CC=48)<sup>§</sup> | 55.00<br>(29.08) | 33.30<br>(27.06) | 85.83<br>(22.63) | 95.90<br>(6.12) | *F(3,87.47)=110.64, **p<0.001**, $\omega^2=0.54$ |
| SF-36 Role Limitations – Physical: M (SD) (FS=49; CC=48)<sup>§</sup> | 16.33<br>(30.0) | 14.00<br>(25.83) | 71.88<br>(39.82) | 90.50<br>(24.16) | *F(3,105.38)=103.07, **p<0.001**, $\omega^2=0.55$ |
| SF-36 Role Limitations – Emotional: M (SD) (FMS=48; CC=48)<sup>§</sup> | 38.00<br>(44.68) | 50.69<br>(45.61) | 45.14<br>(40.38) | 86.67<br>(29.36) | *F(3,104.31)=20.08, **p<0.001**, $\omega^2=0.17$ |
| SF-36 Emotional Wellbeing: M (SD) (CC=48)<sup>§</sup> | 52.88<br>(19.08) | 58.24<br>(22.90) | 51.75<br>(20.67) | 78.72<br>(11.55) | *F(3,102.96)=36.94, **p<0.001**, $\omega^2=0.24$ |
| SF-36 Pain: M (SD) (CC=48)<sup>§</sup> | 41.50<br>(23.55) | 36.55<br>(22.59) | 64.06<br>(25.15) | 82.50<br>(17.65) | *F(3,106.43)=55.06, **p<0.001**, $\omega^2=0.40$ |
| SF-36 Social Functioning: M (SD) (FS=49; FMS=49; CC=48)<sup>§</sup> | 39.29<br>(25.77) | 39.03<br>(26.35) | 62.50<br>(29.29) | 90.80<br>(14.46) | *F(3,100.91)=80.24, **p<0.001**, $\omega^2=0.43$ |
| SF-36 Energy: M (SD) (FS=46; FMS=45; HC=49; CC=48)<sup>§</sup> | 29.57<br>(19.11) | 25.23<br>(15.70) | 34.38<br>(19.91) | 60.71<br>(14.54) | F(3,184)=39.95, **p<0.001**, $\eta^2=0.41$ |
| *SF-36 General Health: M (SD)*<br>(CC=48)<sup>§</sup> | 36.90<br>(20.63) | 33.30<br>(22.17) | 51.88<br>(21.99) | 75.10<br>(16.74) | F(3,194)=42.98, **p<0.001**,<br>$\eta^2=0.40$ |
| **Work and Social Adjustment**<br>**Scale: M (SD) (CC=48)<sup>§</sup>** | 25.64<br>(9.42) | 26.60<br>(7.97) | 17.50<br>(10.49) | 2.62<br>(4.44) | *F(3,100.47)=166.89,<br>**p<0.001**, $\omega^2=0.57$ |

**Key:** CC= clinical control; FMS= functional motor symptoms; FS=functional seizures; HC=healthy control; M=mean; SD=standard deviation
\*Welch's ANOVA <sup>§</sup>ANOVA included only FS/FMS/CC groups due to floor effects in HC group <sup>§</sup>Sample size deviated due to missing data

**Supplementary Table 6.**
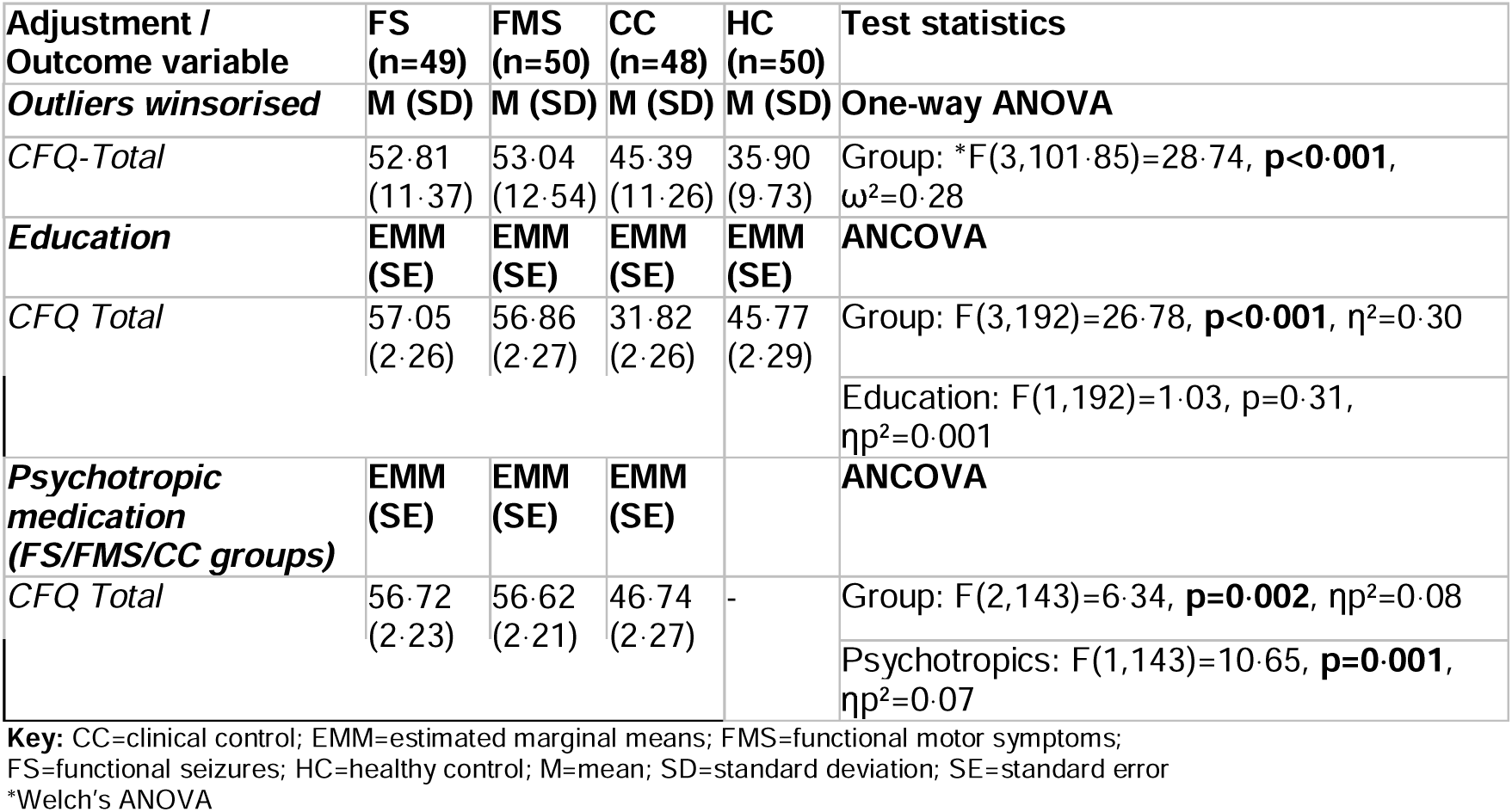
Cognitive Failures Questionnaire sensitivity analyses.

**Supplementary Table 7.**
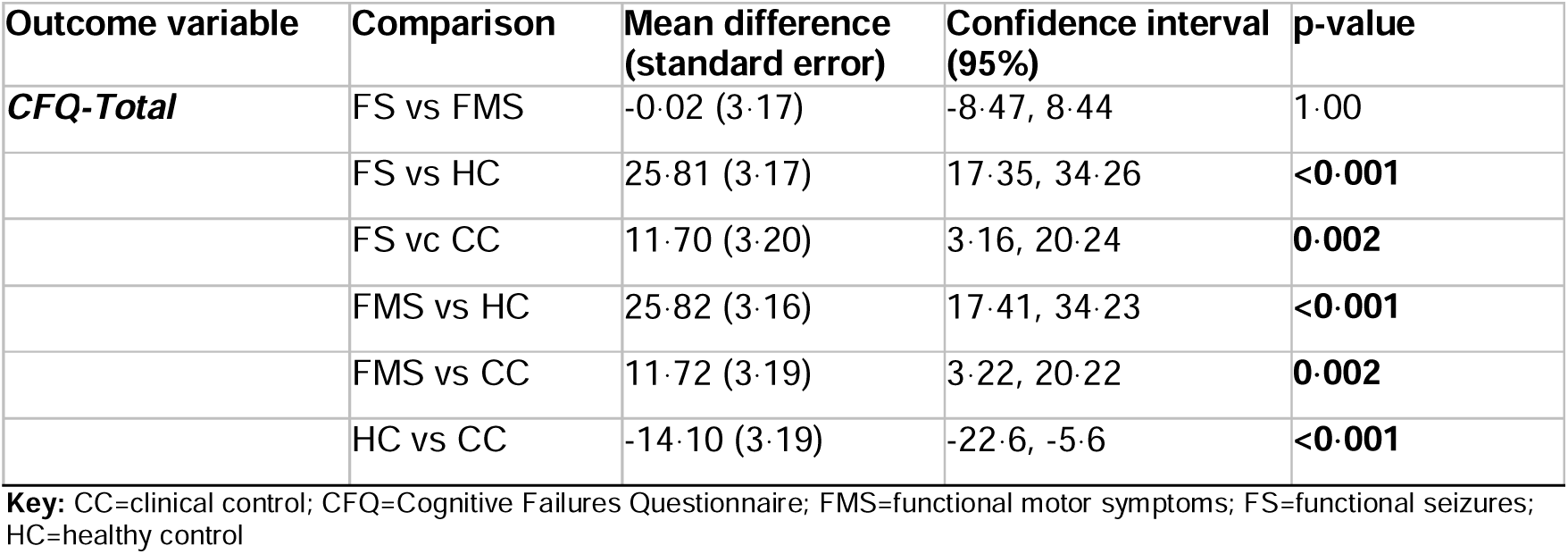
Cognitive Failures Questionnaire Bonferroni post-hoc tests.

| Outcome variable | Comparison | Mean difference (standard error) | Confidence interval (95%) | p-value |
| --- | --- | --- | --- | --- |
| **CFQ-Total** | FS vs FMS | -0.02 (3.17) | -8.47, 8.44 | 1.00 |
|  | FS vs HC | 25.81 (3.17) | 17.35, 34.26 | **<0.001** |
|  | FS vs CC | 11.70 (3.20) | 3.16, 20.24 | **0.002** |
|  | FMS vs HC | 25.82 (3.16) | 17.41, 34.23 | **<0.001** |
|  | FMS vs CC | 11.72 (3.19) | 3.22, 20.22 | **0.002** |
|  | HC vs CC | -14.10 (3.19) | -22.6, -5.6 | **<0.001** |

**Key:** CC=clinical control; CFQ=Cognitive Failures Questionnaire; FMS=functional motor symptoms; FS=functional seizures; HC=healthy control

**Supplementary Table 8.** WASI-II post-hoc tests.

| Test Outcome | Comparison | Mean difference (standard error) | Confidence interval (95%) | p-value |
| --- | --- | --- | --- | --- |
| **WASI-II Full Scale IQ<sup>#</sup>** | FS vs FMS | -2.04 (2.15) | -7.67, 3.59 | 0.78 |
|  | FS vs HC | -3.84 (1.68) | -8.25, 0.57 | 0.11 |
|  | FS vs CC | -6.36 (1.88) | -11.26, -1.46 | **0.005** |
|  | FMS vs HC | -1.80 (1.96) | -6.93, 3.33 | 0.79 |
|  | FMS vs CC | -4.32 (2.12) | -9.87, 1.23 | 0.18 |
|  | HC vs CC | -2.52 (1.65) | -6.83, 1.79 | 0.42 |
| **WASI-II Vocabulary T scores<sup>\*</sup>** | FS vs FMS | -2.04 (1.92) | -7.15, 3.07 | 1.00 |
|  | FS vs HC | -3.84 (1.92) | -8.95, 1.27 | 0.28 |
|  | FS vs CC | -6.36 (1.92) | -11.47, -1.25 | **0.006** |
|  | FMS vs HC | -1.80 (1.92) | -6.91, 3.31 | 1.00 |
|  | FMS vs CC | -4.32 (1.92) | -9.43, 0.79 | 0.15 |
|  | HC vs CC | -2.52 (1.92) | -7.63, 2.59 | 1.00 |
| **WASI-II Matrix Reasoning T scores<sup>\*</sup>** | FS vs FMS | 0.96 (1.80) | -3.84, 5.76 | 1.00 |
|  | FS vs HC | -6.14 (1.80) | -10.94, -1.34 | **0.005** |
|  | FS vs CC | -6.12 (1.80) | -10.92, -1.32 | **0.005** |
|  | FMS vs HC | -7.10 (1.80) | -11.90, -2.30 | **<0.001** |
|  | FMS vs CC | -7.08 (1.80) | -11.88, -2.28 | **<0.001** |
|  | HC vs CC | 0.02 (1.80) | -4.78, 4.82 | 1.00 |

**Key:** CC=clinical control; CFQ=Cognitive Failures Questionnaire; FMS=functional motor symptoms; FS=functional seizures; HC=healthy control; IQ=intelligence quotient; WASI-II=Wechsler Abbreviated Scale of Intelligence – Second edition
<sup>\*</sup>Bonferroni; <sup>#</sup>Games-Howell

**Supplementary Table 9.**
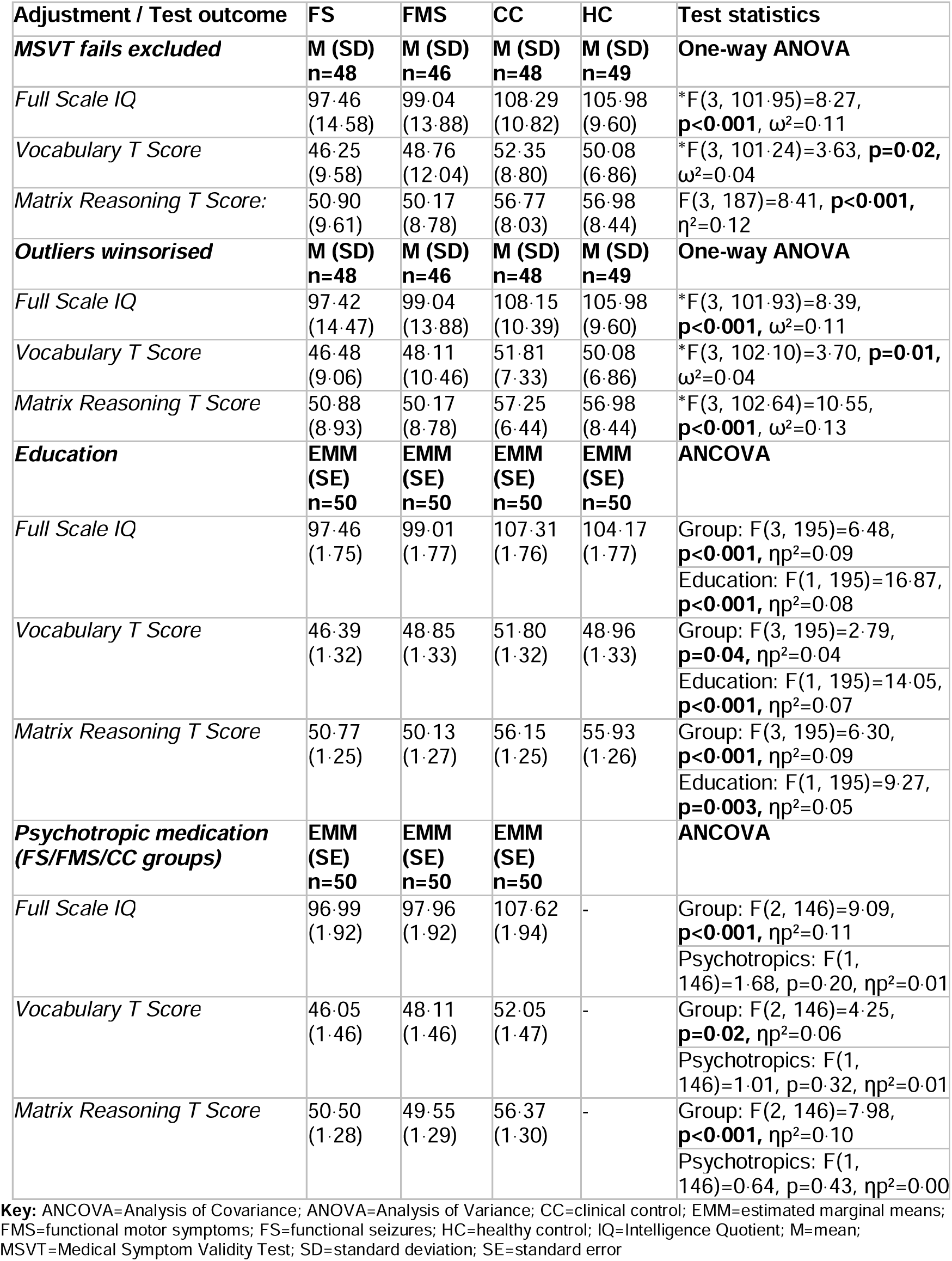
WASI-II sensitivity analyses.

**Supplementary Table 10.**
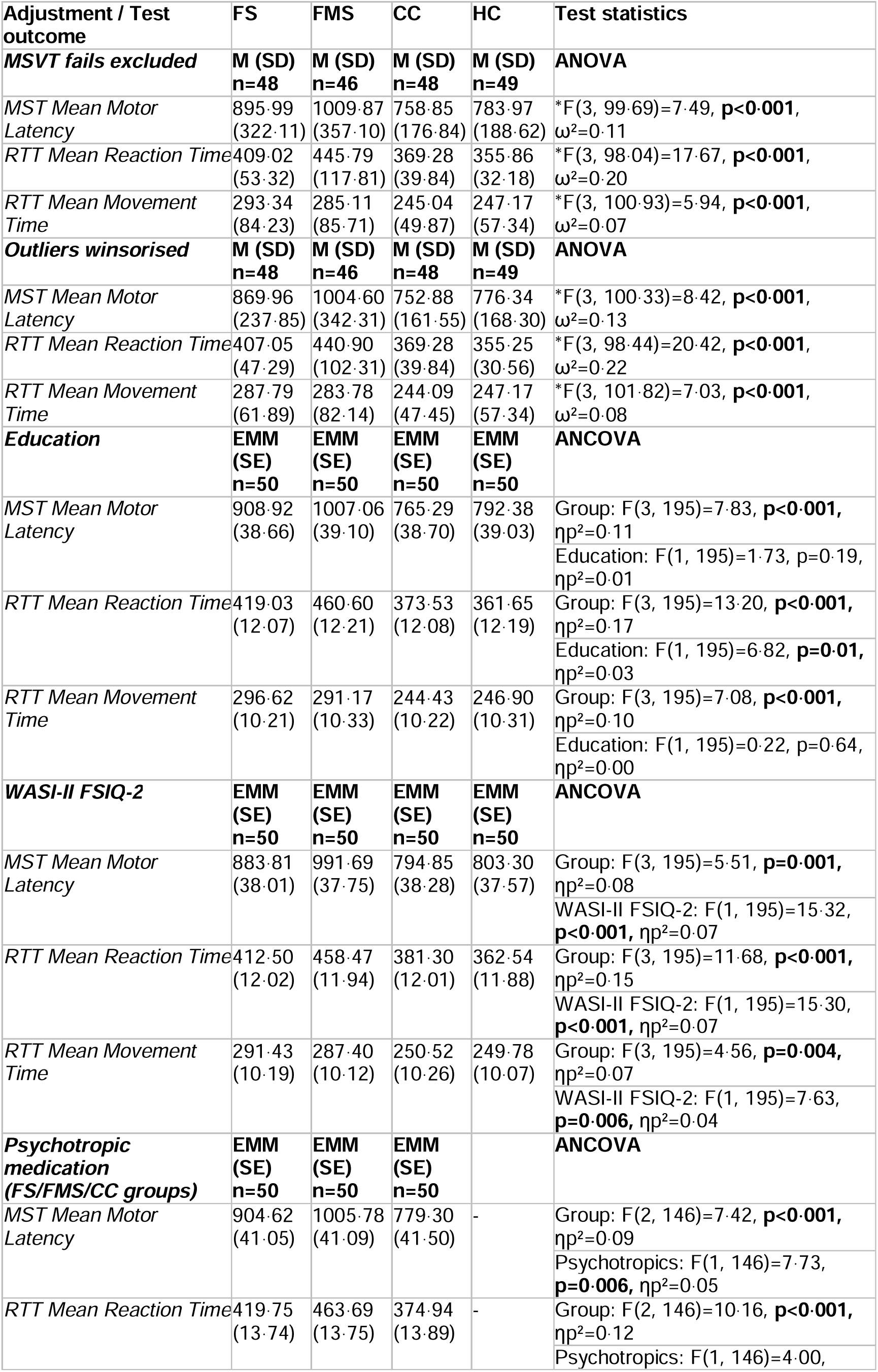
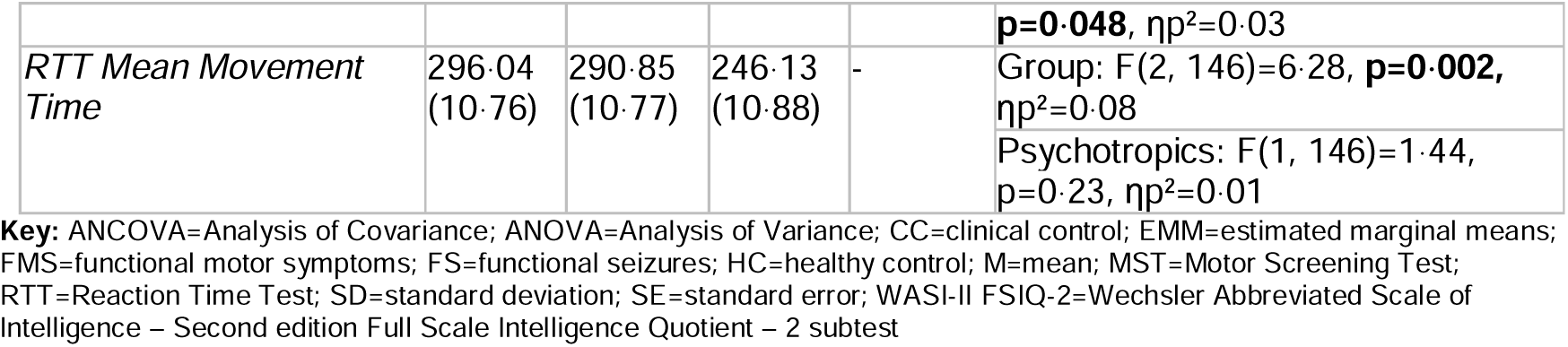
Motor Screening and Reaction Time Test sensitivity analyses.

**Supplementary Table 11.** Motor screening and reaction time Games-Howell post-hoc tests.

| Test outcome | Comparison | Mean difference (standard error) | Confidence interval (95%) | p-value |
| --- | --- | --- | --- | --- |
| ***MST Mean Motor Latency*** | FS vs FMS | -102.49 (67.96) | -280.15, 75.16 | 0.44 |
|  | FS vs HC | 129.60 (53.29) | -10.31, 269.52 | 0.08 |
|  | FS vs CC | 153.43 (52.71) | 14.95, 291.91 | **0.02** |
|  | FMS vs HC | 232.10 (56.33) | 84.06, 380.13 | **<0.001** |
|  | FMS vs CC | 255.92 (55.79) | 109.23, 402.61 | **<0.001** |
|  | HC vs CC | 23.82 (36.51) | -71.60, 119.25 | 0.91 |
| ***RTT Mean Reaction Time*** | FS vs FMS | -44.26 (23.26) | -105.31, 16.78 | 0.24 |
|  | FS vs HC | 65.47 (12.74) | 31.86, 99.08 | **<0.001** |
|  | FS vs CC | 51.57 (13.22) | 16.79, 86.36 | **0.001** |
|  | FMS vs HC | 109.73 (20.50) | 55.39, 164.08 | **<0.001** |
|  | FMS vs CC | 95.84 (20.80) | 40.78, 150.89 | **<0.001** |
|  | HC vs CC | -13.90 (7.33) | -33.08, 5.28 | 0.24 |
| ***RTT Mean Movement Time*** | FS vs FMS | 5.04 (17.19) | -39.89, 49.97 | 0.99 |
|  | FS vs HC | 50.93 (14.55) | 12.80, 89.07 | **0.004** |
|  | FS vs CC | 53.10 (14.04) | 16.25, 89.95 | **0.002** |
|  | FMS vs HC | 45.89 (14.65) | 7.49, 84.29 | **0.01** |
|  | FMS vs CC | 48.06 (14.15) | 10.93, 85.18 | **0.006** |
|  | HC vs CC | 2.166 (10.79) | -26.04, 30.37 | 0.10 |

**Key:** CC=clinical control; FMS=functional motor symptoms; FS=functional seizures; HC=healthy control; MST=Motor Screening Test; RTT=Reaction Time Test

**Supplementary Table 12.**
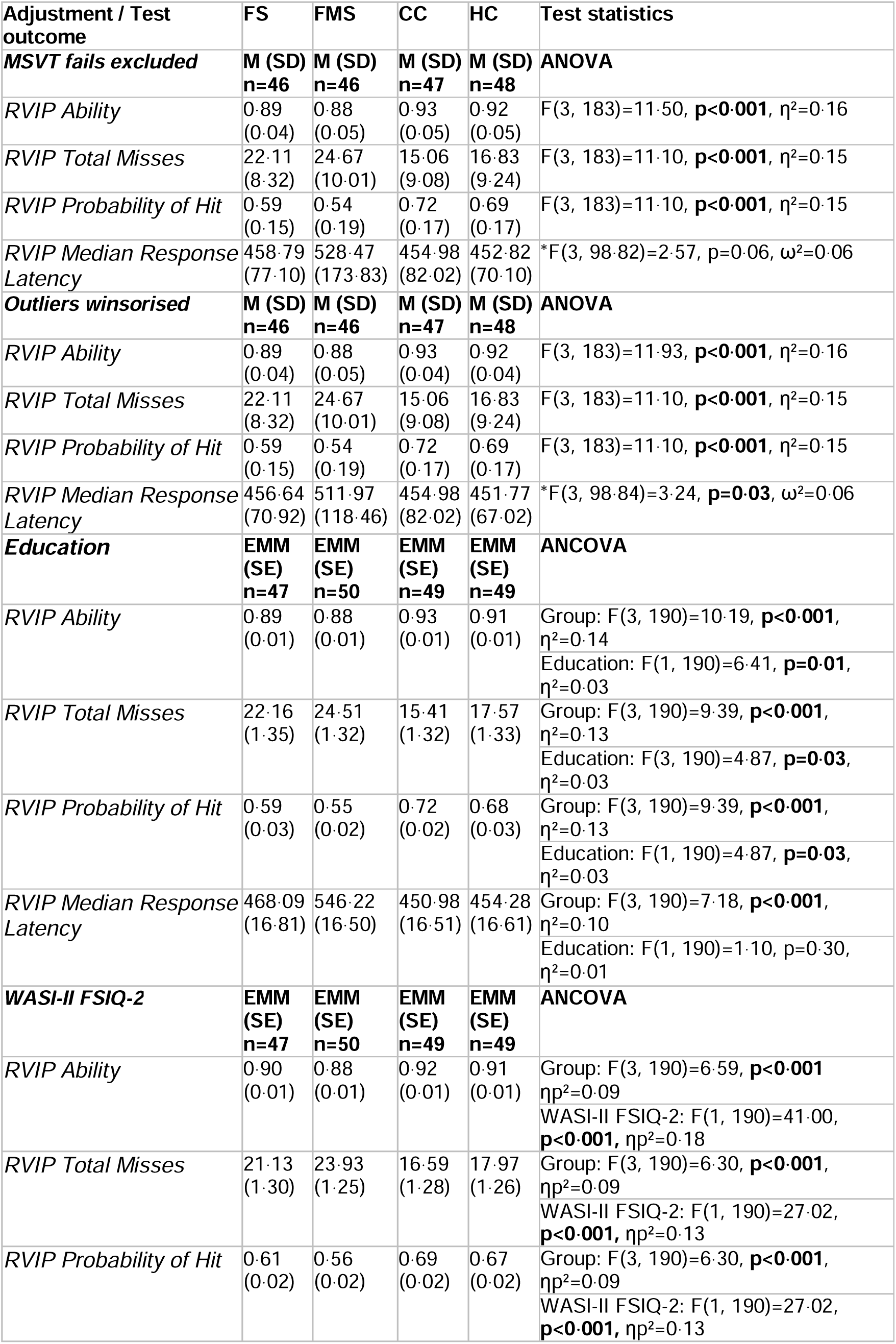
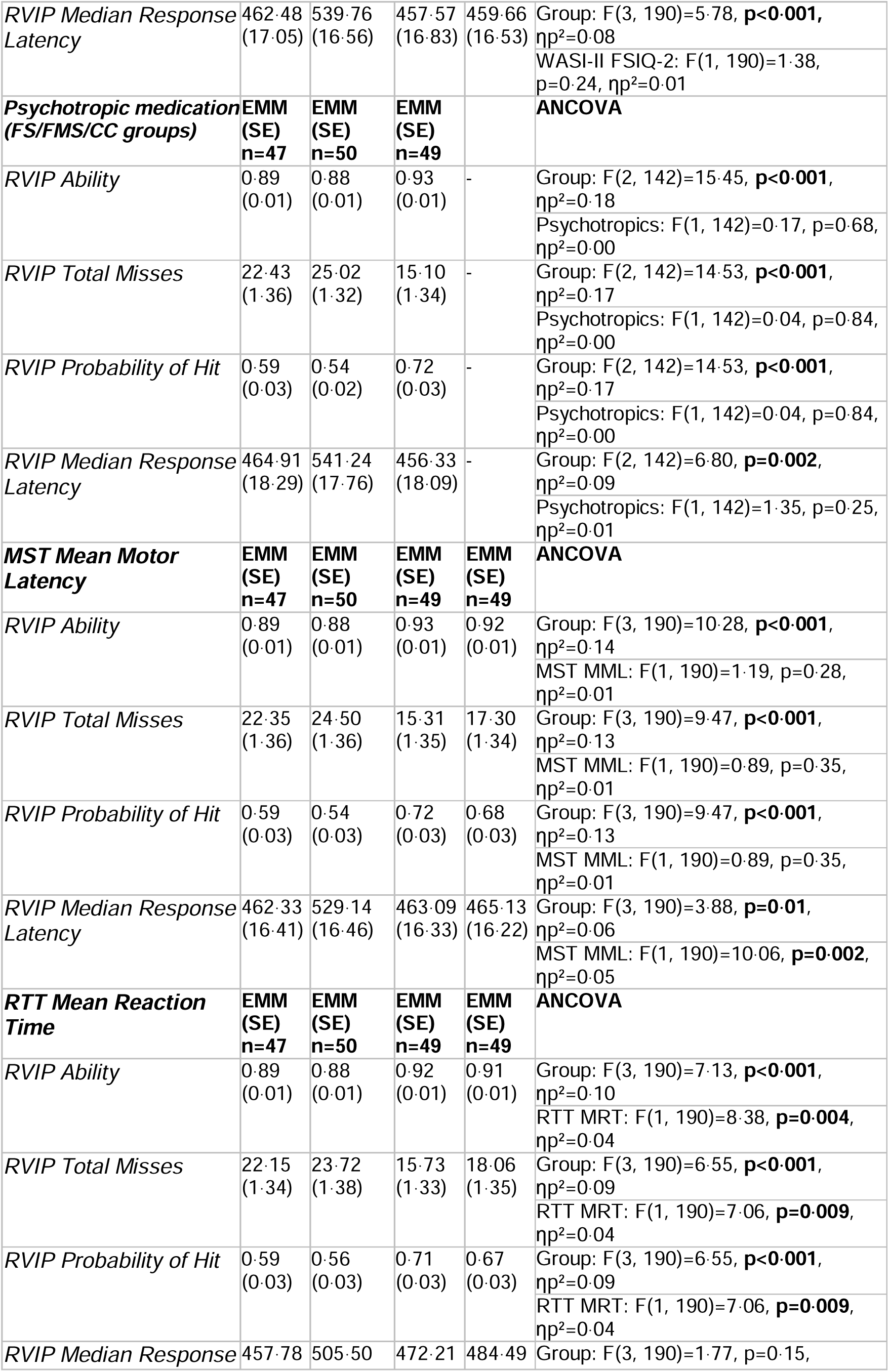
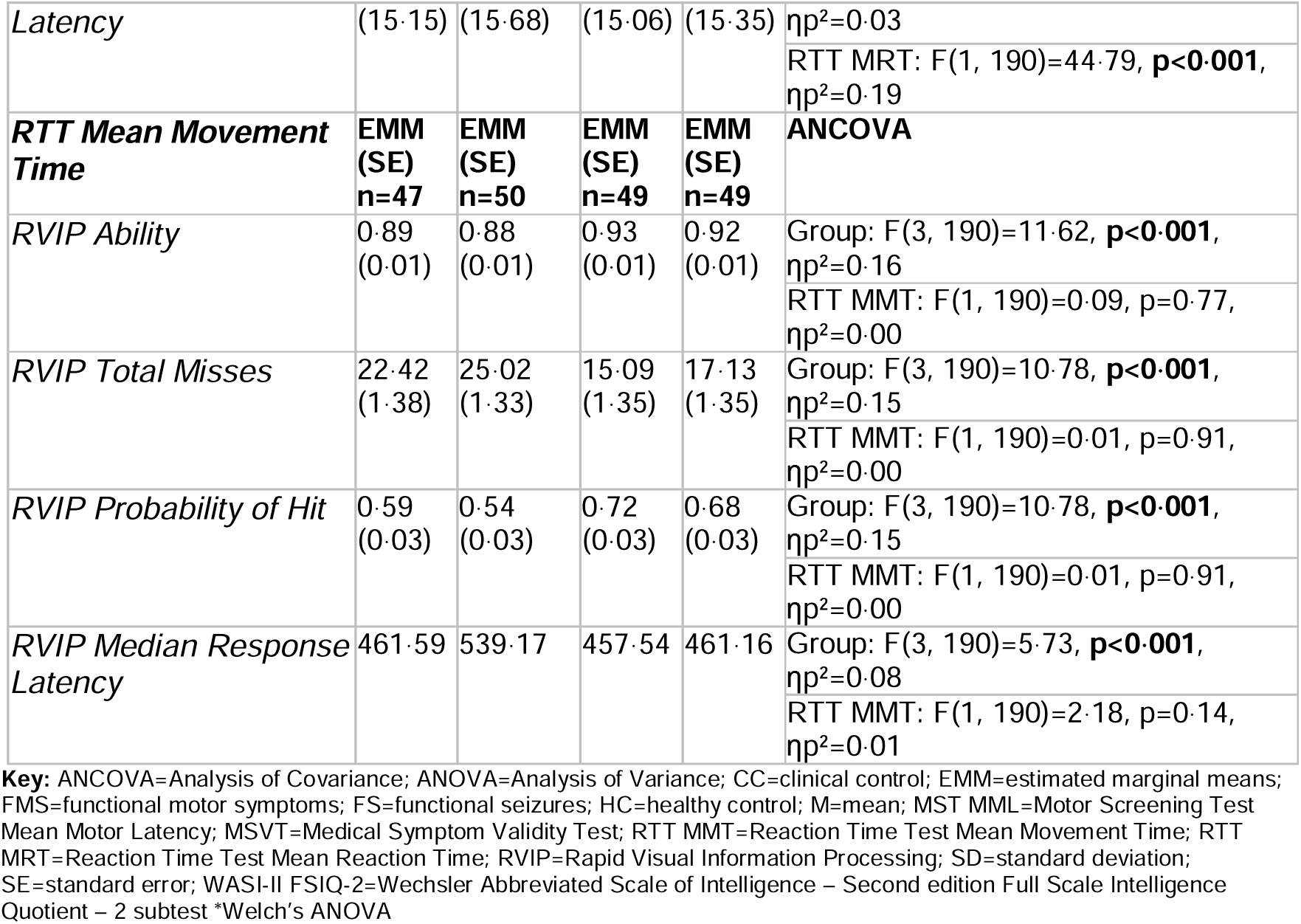
Rapid Visual Information Processing sensitivity analyses.

**Supplementary Table 13.** Rapid Visual Information Processing Bonferroni post-hoc tests.

| Test outcome | Comparison | Mean difference (standard error) | Confidence interval (95%) | p-value |
| --- | --- | --- | --- | --- |
| ***RVIP Ability*** | FS vs FMS | 0.01 (0.01) | -0.01, 0.04 | 0.92 |
|  | FS vs HC | -0.03 (0.01) | -0.05, -0.00 | **0.02** |
|  | FS vs CC | -0.04 (0.01) | -0.06, -0.01 | **<0.001** |
|  | FMS vs HC | -0.04 (0.01) | -0.07, -0.02 | **<0.001** |
|  | FMS vs CC | -0.05 (0.01) | -0.08, -0.03 | **<0.001** |
|  | HC vs CC | -0.01 (0.01) | -0.04, 0.01 | 1.00 |
| ***RVIP Total Misses*** | FS vs FMS | -2.59 (1.89) | -7.62, 2.43 | 1.00 |
|  | FS vs HC | 5.35 (1.90) | 0.29, 10.40 | **0.03** |
|  | FS vs CC | 7.39 (1.90) | 2.33, 12.44 | **<0.001** |
|  | FMS vs HC | 7.94 (1.90) | 2.96, 12.91 | **<0.001** |
|  | FMS vs CC | 9.98 (1.90) | 5.01, 14.95 | **<0.001** |
|  | HC vs CC | 2.04 (1.90) | -2.96, 7.04 | 1.00 |
| ***RVIP Probability of Hit*** | FS vs FMS | 0.05 (0.03) | -0.05, 0.14 | 1.00 |
|  | FS vs HC | -0.10 (0.04) | -0.19, -0.01 | **0.03** |
|  | FS vs CC | -0.14 (0.04) | -0.23, -0.04 | **<0.001** |
|  | FMS vs HC | -0.15 (0.03) | -0.24, -0.05 | **<0.001** |
|  | FMS vs CC | -0.18 (0.03) | -0.28, -0.09 | **<0.001** |
|  | HC vs CC | -0.04 (0.03) | -0.13, 0.05 | 1.00 |

**Key:** CC=clinical control; FMS=functional motor symptoms; FS=functional seizures; HC=healthy control; RVIP=Rapid Visual Information Processing

**Supplementary Table 14.**
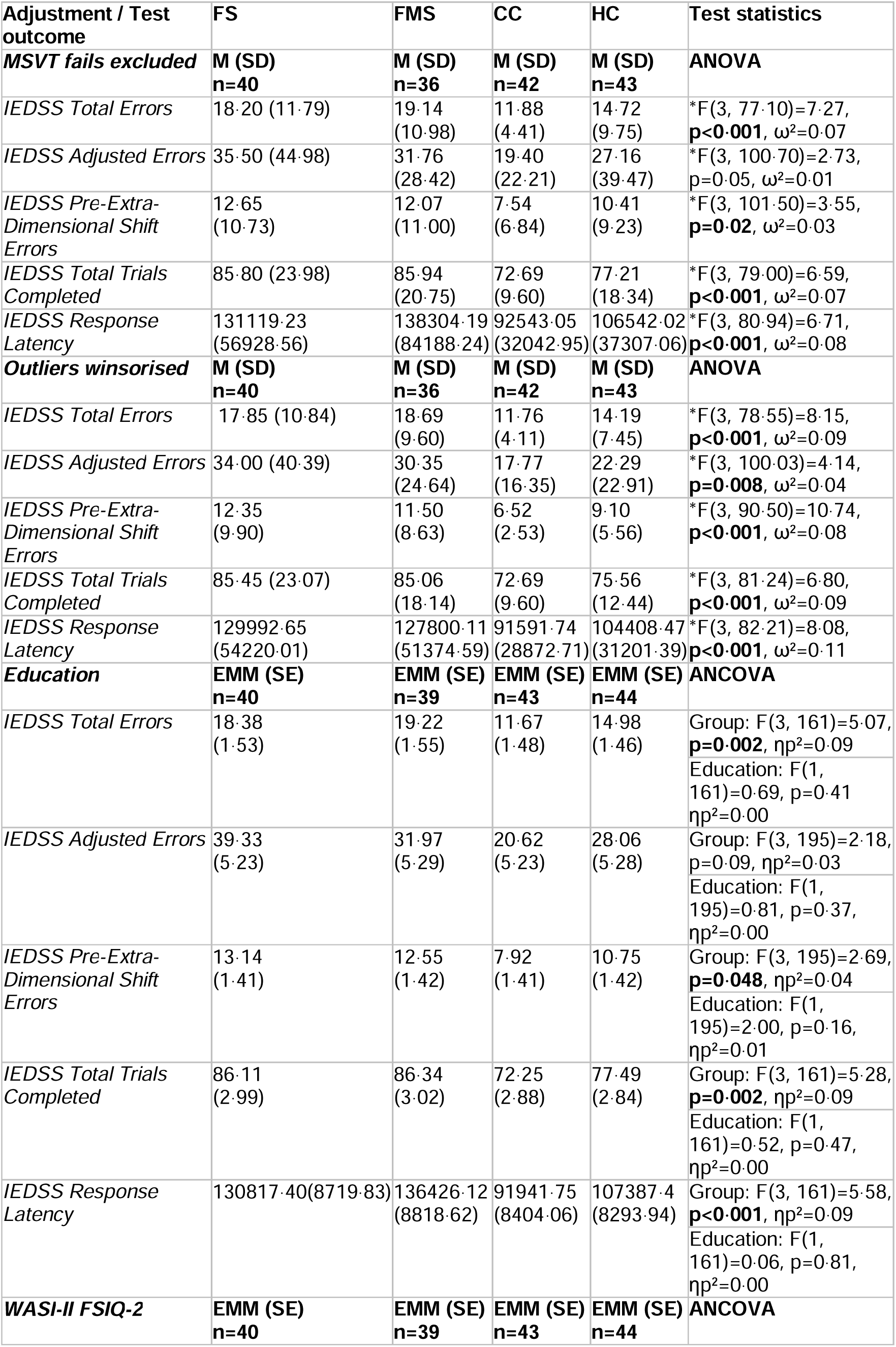
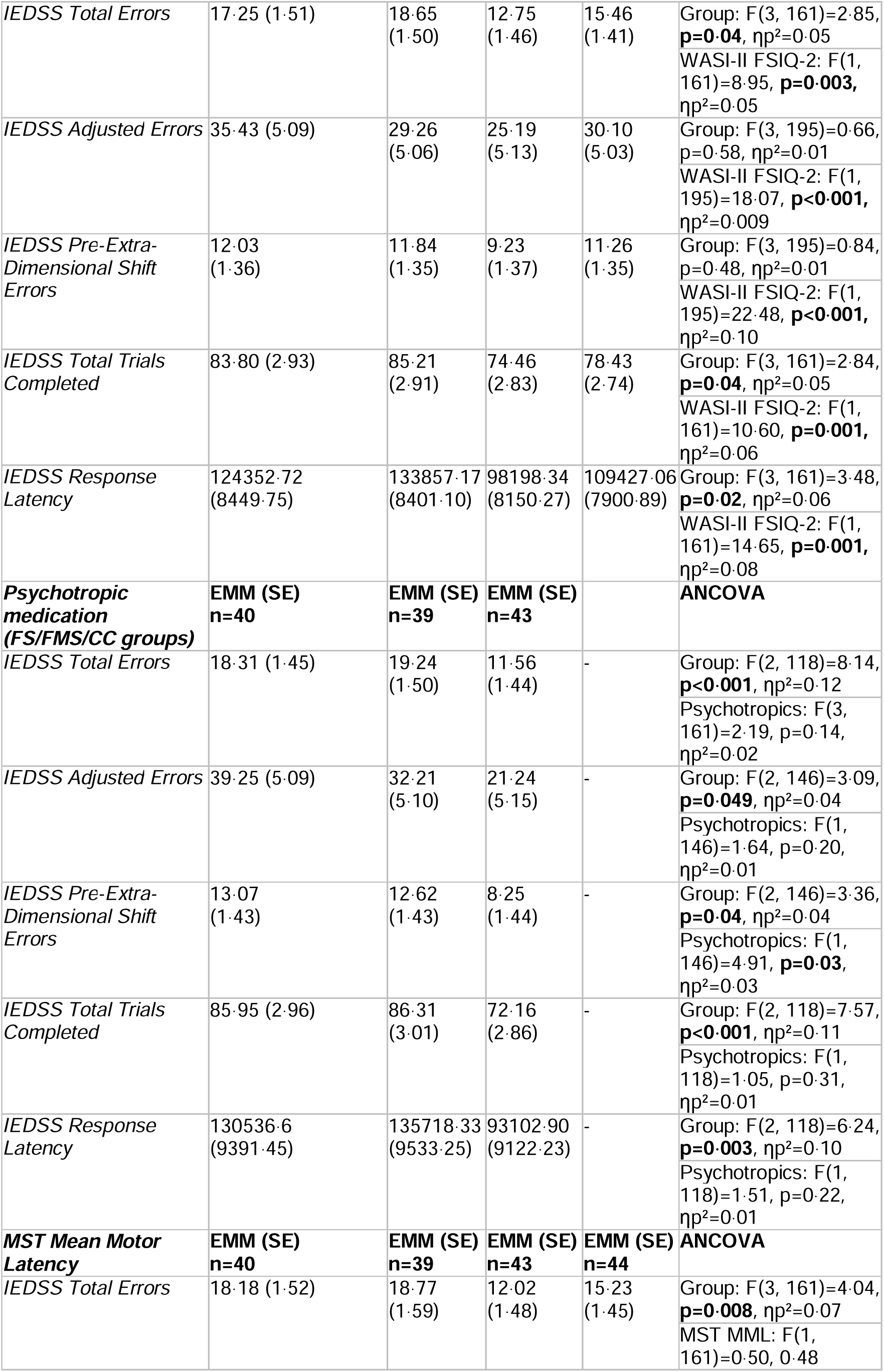
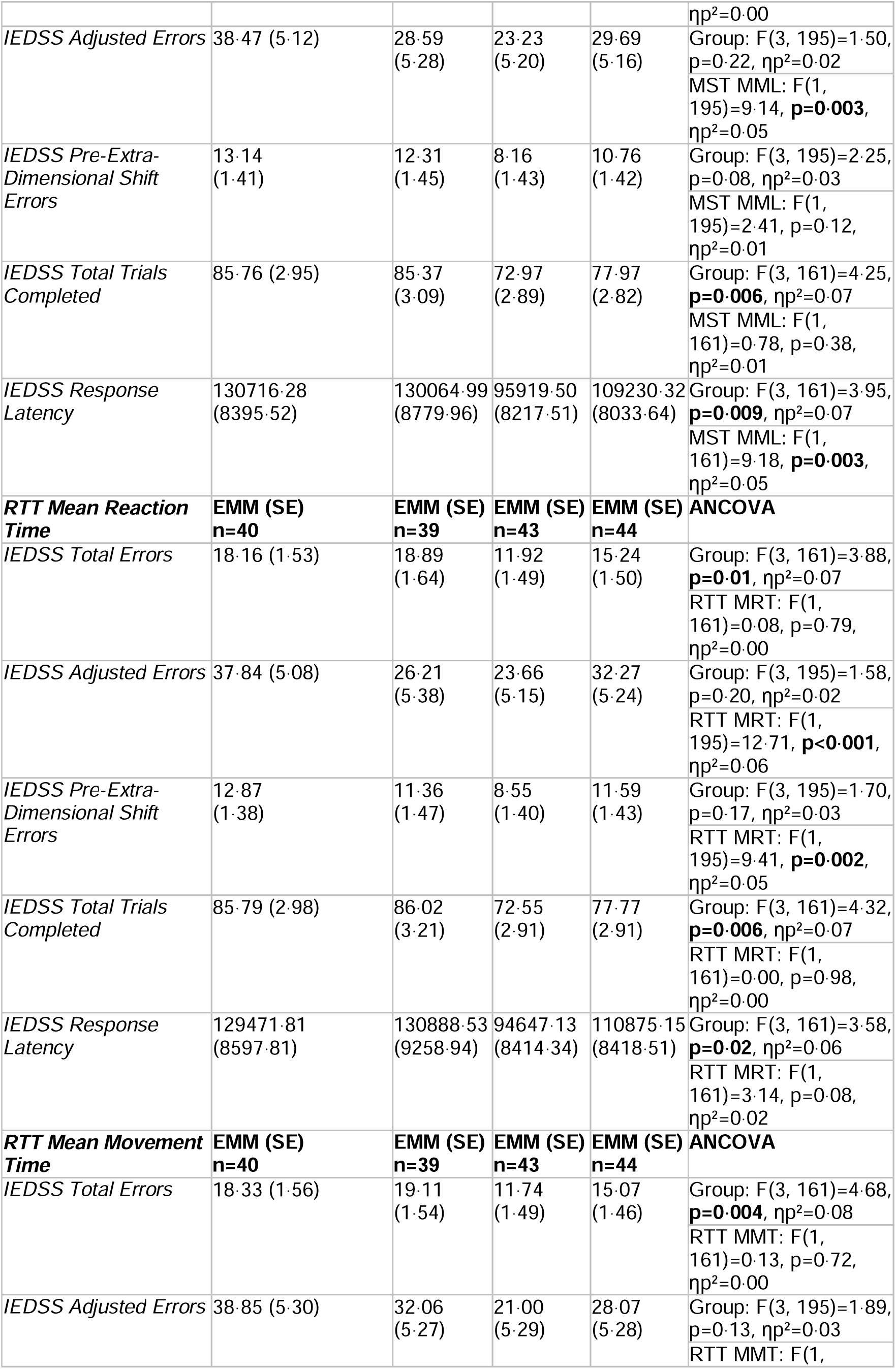
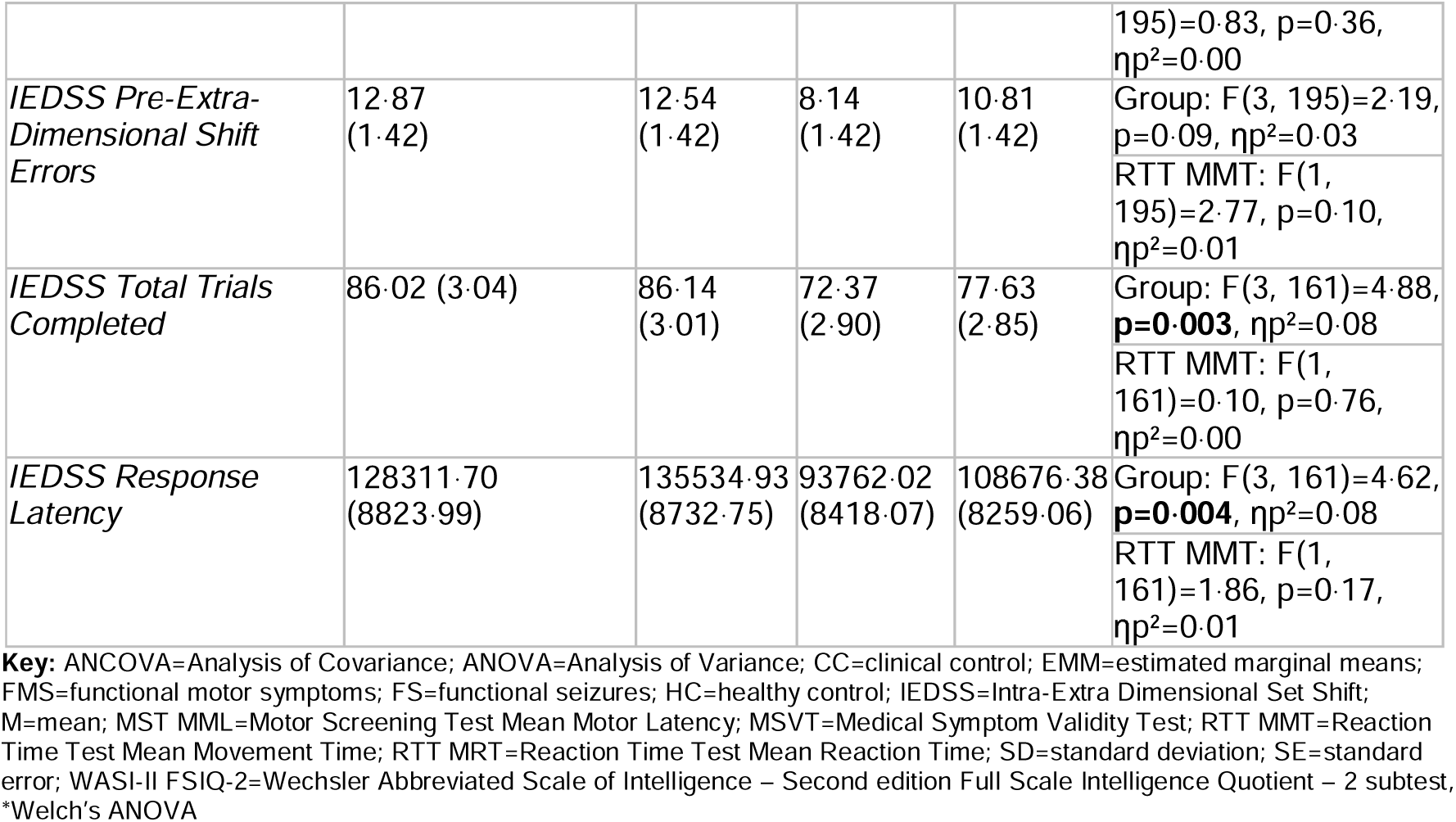
Intra-Extra Dimensional Set-Shift sensitivity analyses.

**Supplementary Table 15.** Intra-Extra Dimensional Set Shift post-hoc tests.

| Test outcome | Comparison | Mean difference (standard error) | Confidence interval (95%) | p-value |
| --- | --- | --- | --- | --- |
| ***IEDSS Total Errors\**** | FS vs FMS | -0.85 (2.53) | -7.48, 5.78 | 0.99 |
|  | FS vs HC | 3.06 (2.40) | -3.24, 9.36 | 0.58 |
|  | FS vs CC | 6.36 (1.98) | 1.10, 11.63 | **0.01** |
|  | FMS vs HC | 3.92 (2.28) | -2.06, 9.89 | 0.32 |
|  | FMS vs CC | 7.21 (1.83) | 2.35, 12.08 | **0.001** |
|  | HC vs CC | 3.30 (1.65) | -1.06, 7.66 | 0.20 |
| ***IEDSS Total Trials Completed\**** | FS vs FMS | -0.25 (4.98) | -13.33, 12.83 | 1.00 |
|  | FS vs HC | 8.05 (4.70) | -4.32, 20.42 | 0.33 |
|  | FS vs CC | 13.27 (4.06) | 2.47, 24.06 | **0.01** |
|  | FMS vs HC | 8.30 (4.26) | -2.89, 19.49 | 0.22 |
|  | FMS vs CC | 13.52 (3.54) | 4.13, 22.91 | **0.002** |
|  | HC vs CC | 5.22 (3.14) | -3.07, 13.50 | 0.35 |
| ***IEDSS Response Latency\**** | FS vs FMS | -5591.37 (15816.16) | -47247.62, 36064.89 | 0.99 |
|  | FS vs HC | 23983.91 (10595.76) | -3943.68, 51911.49 | 0.12 |
|  | FS vs CC | 39458.25 (10252.27) | 12374.75, 66541.74 | **0.002** |
|  | FMS vs HC | 29575.27 (14155.49) | -8000.12, 67150.66 | 0.17 |
|  | FMS vs CC | 45049.61 (13900.25) | 8074.83, 82024.40 | **0.01** |
|  | HC vs CC | 15474.34 (7438.82) | -4025.15, 34973.83 | 0.17 |

**Key:** CC=clinical control; FMS=functional motor symptoms; FS=functional seizures; HC=healthy control; IEDSS=Intra-Extra Dimensional Set Shift, \*Games-Howell

**Supplementary Table 16.** Spatial Span Test uncorrected results.

|  | **FS**<br>**(n=50)** | **FMS**<br>**(n=50)** | **CC**<br>**(n=50)** | **HC**<br>**(n=50)** | **ANOVA statistics** |
| --- | --- | --- | --- | --- | --- |
| *SSP Forward Span Length: M (SD)* | 6.18<br>(1.17) | 6.16<br>(1.39) | 6.74<br>(1.43) | 6.78<br>(1.45) | F(3, 196)=3.13, **p=0.03**, $\eta^2=0.05$ |
| *SSP Forward Errors: M (SD)* | 16.46<br>(6.27) | 15.72<br>(9.84) | 14.0<br>(7.23) | 13.34<br>(5.31) | F(3, 196)=1.95, p=0.122, $\eta^2=0.03$ |
| *SSP Reverse Span Length: M (SD)* | 5.62<br>(1.37) | 5.60<br>(1.21) | 6.60<br>(1.34) | 6.30<br>(1.42) | F(3, 196)=7.00, **p<0.001**, $\eta^2=0.10$ |
| *SSP Reverse Errors: M (SD)* | 12.30<br>(5.02) | 13.06<br>(5.01) | 13.66<br>(5.84) | 12.20<br>(5.80) | F(3, 196)=0.80, p=0.50, $\eta^2=0.01$ |

**Key:** CC=clinical controls; FS=functional seizures; FMS=functional motor symptoms; HC=healthy controls; M=mean; ms=milliseconds; SD=standard deviation †Welch's ANOVA

**Supplementary Table 17.** Spatial Span Test sensitivity analyses.

| Adjustment / Test outcome<sup>B</sup> | FS | FMS | CC | HC | Test statistics |
| --- | --- | --- | --- | --- | --- |
| **MSVT fails excluded** | **M (SD)**<br>**n=48** | **M (SD)**<br>**n=46** | **M (SD)**<br>**n=48** | **M (SD)**<br>**n=49** | **ANOVA** |
| *SSP Forward Span Length* | 6.23<br>(1.17) | 6.33<br>(1.32) | 6.77<br>(1.39) | 6.82<br>(1.44) | F(3, 187)=2.45, p=0.07,<br>η<sup>2</sup>=0.04 |
| *SSP Reverse Span Length* | 5.71<br>(1.32) | 5.70<br>(1.21) | 6.67<br>(1.26) | 6.33<br>(1.42) | F(3, 187)=6.41, **p<0.001**,<br>η<sup>2</sup>=0.09 |
| **Education** | **EMM**<br>**(SE)**<br>**n=50** | **EMM**<br>**(SE)**<br>**n=50** | **EMM**<br>**(SE)**<br>**n=50** | **EMM**<br>**(SE)**<br>**n=50** | **ANCOVA** |
| *SSP Forward Span Length* | 6.19<br>(0.19) | 6.18<br>(0.20) | 6.73<br>(0.19) | 6.76<br>(0.20) | Group: F(3, 195)=2.63,<br>p=0.05, ηp<sup>2</sup>=0.04<br>Education: F(1, 195)=0.29,<br>p=0.59, ηp<sup>2</sup>=0.00 |
| *SSP Reverse Span Length* | 5.64<br>(0.19) | 5.63<br>(0.19) | 6.58<br>(0.19) | 6.27<br>(0.19) | Group: F(3, 195)=5.70,<br>**p<0.001**, ηp<sup>2</sup>=0.08<br>Education: F(1, 195)=0.82,<br>p=0.37, ηp<sup>2</sup>=0.00 |
| **WASI-II FSIQ-2** | **EMM**<br>**(SE)**<br>**n=50** | **EMM**<br>**(SE)**<br>**n=50** | **EMM**<br>**(SE)**<br>**n=50** | **EMM**<br>**(SE)**<br>**n=50** | **ANCOVA** |
| *SSP Forward Span Length* | 6.38<br>(0.18) | 6.32<br>(0.18) | 6.51<br>(0.19) | 6.65<br>(0.18) | Group: F(3, 195)=0.61, p=0.61<br>ηp<sup>2</sup>=0.009<br>WASI-II FSIQ-2: F(1,<br>195)=28.75, **p<0.001**,<br>ηp<sup>2</sup>=0.13 |
| *SSP Reverse Span Length* | 5.82<br>(0.18) | 5.76<br>(0.18) | 6.37<br>(0.18) | 6.17<br>(0.18) | Group: F(3, 195)=2.36, p=0.07<br>ηp<sup>2</sup>=0.04<br>WASI-II FSIQ-2: F(1,<br>195)=30.36, **p<0.001**,<br>ηp<sup>2</sup>=0.14 |
| **Psychotropic medication**<br>**(FS/FMS/CC groups)** | **EMM**<br>**(SE)**<br>**n=50** | **EMM**<br>**(SE)**<br>**n=50** | **EMM**<br>**(SE)**<br>**n=50** |  | **ANCOVA** |
| *SSP Forward Span Length* | 6.20<br>(0.19) | 6.18<br>(0.19) | 6.70<br>(0.19) | - | Group: F(2, 146)=2.33, p=0.10<br>ηp<sup>2</sup>=0.03<br>Psychotropics: F(1, 146)=1.70,<br>p=0.19, ηp<sup>2</sup>=0.01 |
| *SSP Reverse Span Length* | 5.64<br>(0.19) | 5.62<br>(0.19) | 6.57<br>(0.19) | - | Group: F(2, 146)=8.25,<br>**p<0.001**, ηp<sup>2</sup>=0.10<br>Psychotropics: F(1, 146)=1.16,<br>p=0.28, ηp<sup>2</sup>=0.01 |
| **MST Mean Motor Latency** | **EMM**<br>**(SE)**<br>**n=50** | **EMM**<br>**(SE)**<br>**n=50** | **EMM**<br>**(SE)**<br>**n=50** | **EMM**<br>**(SE)**<br>**n=50** | **ANCOVA** |
| *SSP Forward Span Length* | 6.22<br>(0.19) | 6.29<br>(0.20) | 6.64<br>(0.19) | 6.71<br>(0.19) | Group: F(3, 195)=1.50, p=0.22<br>ηp<sup>2</sup>=0.02<br>MST MML: F(1, 195)=6.31,<br>**p=0.01**, ηp<sup>2</sup>=0.03 |
| *SSP Reverse Span Length* | 5.65<br>(0.19) | 5.71<br>(0.19) | 6.52<br>(0.19) | 6.24<br>(0.19) | Group: F(3, 195)=4.46,<br>**p=0.005**, ηp<sup>2</sup>=0.06<br>MST MML: F(1, 195)=4.80,<br>**p=0.03**, ηp<sup>2</sup>=0.02 |
| **RTT Mean Reaction Time** | **EMM**<br>**(SE)**<br>**n=50** | **EMM**<br>**(SE)**<br>**n=50** | **EMM**<br>**(SE)**<br>**n=50** | **EMM**<br>**(SE)**<br>**n=50** |  |
| *SSP Forward Span Length* | 6.26<br>(0.19) | 6.43<br>(0.19) | 6.59<br>(0.19) | 6.57<br>(0.19) | Group: F(3, 195)=0.63,<br>p=0.60, $\eta^2$=0.01<br>RTT MRT: F(1, 195)=16.30,<br>**p<0.001**, $\eta^2$=0.08 |
| *SSP Reverse Span Length* | 5.70<br>(0.18) | 5.87<br>(0.19) | 6.46<br>(0.19) | 6.10<br>(0.19) | Group: F(3, 195)=2.92,<br>**p=0.04**, $\eta^2$=0.04<br>RTT MRT: F(1, 195)=16.56,<br>**p<0.001**, $\eta^2$=0.08 |
| ***RTT Mean Movement Time*** | **EMM**<br>**(SE)**<br>**n=50** | **EMM**<br>**(SE)**<br>**n=50** | **EMM**<br>**(SE)**<br>**n=50** | **EMM**<br>**(SE)**<br>**n=50** |  |
| *SSP Forward Span Length* | 6.27<br>(0.19) | 6.23<br>(0.19) | 6.66<br>(0.19) | 6.71<br>(0.19) | Group: F(3, 195)=1.55,<br>p=0.20, $\eta^2$=0.02<br>RTT MMT: F(1, 195)=5.65,<br>**p=0.02**, $\eta^2$=0.03 |
| *SSP Reverse Span Length* | 5.70<br>(0.19) | 5.66<br>(0.19) | 6.53<br>(0.19) | 6.24<br>(0.19) | Group: F(3, 195)=4.60,<br>**p=0.004**, $\eta^2$=0.07<br>RTT MMT: F(1, 195)=4.34,<br>**p=0.04**, $\eta^2$=0.02 |

**Key:** ANCOVA=Analysis of Covariance; ANOVA=Analysis of Variance; CC=clinical control; EMM=estimated marginal means; FMS=functional motor symptoms; FS=functional seizures; HC=healthy control; M=mean; MMT=Mean Movement Time; MST MML=Motor Screening Test Mean Motor Latency; MRT=Mean Reaction Time; MSVT=Medical Symptom Validity Test; RTT=Reaction Time Test; SD=standard deviation; SE=standard error; SSP=Spatial Span Test; WASI-II FSIQ-2=Wechsler Abbreviated Scale of Intelligence – Second edition Full Scale Intelligence Quotient – 2 subtest
<sup>§</sup>No outliers present

**Supplementary Table 18.** Stop Signal Task uncorrected results.

|  | **FS**<br>**(n=49)** | **FMS**<br>**(n=48)** | **CC**<br>**(n=50)** | **HC**<br>**(n=50)** | **ANOVA statistics** |
| --- | --- | --- | --- | --- | --- |
| *Errors Go Trials: M (SD)* | 2.71<br>(4.35) | 2.94<br>(4.22) | 1.56<br>(2.55) | 1.64<br>(2.20) | *F(3,102.91)=2.04, p=0.113,<br>$\omega^2=0.02$ |
| *Errors Stop Trials: M (SD)* | 40.69<br>(4.67) | 42.27<br>(4.00) | 40.86<br>(4.00) | 41.36<br>(5.50) | F(3, 193)=1.03, p=0.38,<br>$\omega^2=0.02$ |
| *Number of Missed Trials: M (SD)* | 6.67<br>(6.08) | 6.19<br>(5.95) | 3.88<br>(3.81) | 5.46<br>(11.28) | F(3,193)=1.37, **p=0.03**, $\eta^2=0.02$ |
| *Stop Signal Reaction Time (ms):*<br>*M (SD)* | 244.02<br>(56.98) | 253.91<br>(58.47) | 229.42<br>(47.93) | 243.24<br>(59.85) | F(3, 193)=1.59, p=0.19,<br>$\eta^2=0.02$ |

**Key:** CC=clinical controls; FS=functional seizures; FMS=functional motor symptoms; HC=healthy controls; M=mean; ms=milliseconds; SD=standard deviation \*Welch's ANOVA

**Supplementary Table 19.** Stop Signal Task sensitivity analyses.

| Adjustment / Test outcome | FS | FMS | CC | HC | Test statistics |
| --- | --- | --- | --- | --- | --- |
| **MSVT fails excluded** | **M (SD)**<br>**n=48** | **M (SD)**<br>**n=45** | **M (SD)**<br>**n=48** | **M (SD)**<br>**n=49** | **ANOVA** |
| *SST Number of Missed Trials* | 6.479<br>(5.99) | 5.96<br>(5.81) | 3.79<br>(3.72) | 5.51<br>(11.39) | F(3, 186)=1.20, p=0.31, $\omega^2=0.02$ |
| **Outliers winsorised** | **M (SD)**<br>**n=48** | **M (SD)**<br>**n=45** | **M (SD)**<br>**n=48** | **M (SD)**<br>**n=49** | **ANOVA** |
| *SST Number of Missed Trials* | 6.31<br>(5.49) | 5.78<br>(5.28) | 3.63<br>(3.32) | 4.41<br>(5.12) | *F(3, 100.25)=3.66, **p=0.02**, $\omega^2=0.03$ |
| **Education** | **EMM (SE)**<br>**n=49** | **EMM (SE)**<br>**n=48** | **EMM (SE)**<br>**n=50** | **EMM (SE)**<br>**n=50** | **ANCOVA** |
| *SST Number of Missed Trials* | 6.65<br>(1.05) | 6.14<br>(1.08) | 3.91<br>(1.05) | 5.51<br>(1.05) | Group: F(3, 192)=1.26, p=0.29, $\eta^2=0.02$<br>Education: F(1, 192)=0.07, p=0.79, $\eta^2=0.00$ |
| **WASI-II FSIQ-2** | **EMM (SE)**<br>**n=49** | **EMM (SE)**<br>**n=48** | **EMM (SE)**<br>**n=50** | **EMM (SE)**<br>**n=50** | **ANCOVA** |
| *SST Number of Missed Trials* | 6.59<br>(1.07) | 6.12<br>(1.08) | 3.98<br>(1.07) | 5.51<br>(1.05) | Group: F(3, 192)=1.07, p=0.37, $\eta^2=0.02$<br>WASI-II FSIQ-2: F(1, 192)=0.16, p=0.69, $\eta^2=0.00$ |
| **Psychotropic medication (FS/FMS/CC groups)** | **EMM (SE)**<br>**n=49** | **EMM (SE)**<br>**n=48** | **EMM (SE)**<br>**n=50** |  | **ANCOVA** |
| *SST Number of Missed Trials* | 6.61<br>(0.77) | 6.10<br>(0.78) | 4.03<br>(0.77) | - | Group: F(2, 143)=3.09, **p=0.049**, $\eta^2=0.04$<br>Psychotropics: F(1, 143)=1.35, p=0.25, $\eta^2=0.01$ |
| **MST Mean Motor Latency** | **EMM (SE)**<br>**n=49** | **EMM (SE)**<br>**n=48** | **EMM (SE)**<br>**n=50** | **EMM (SE)**<br>**n=50** | **ANCOVA** |
| *SST Number of Missed Trials* | 6.54<br>(1.04) | 5.63<br>(1.10) | 4.26<br>(1.05) | 5.75<br>(1.04) | Group: F(3, 192)=0.81, p=0.49, $\eta^2=0.01$<br>MST MML: F(1, 192)=3.43, p=0.07, $\eta^2=0.02$ |
| **RTT Mean Reaction Time** | **EMM (SE)**<br>**n=49** | **EMM (SE)**<br>**n=48** | **EMM (SE)**<br>**n=50** | **EMM (SE)**<br>**n=50** | **ANCOVA** |
| *SST Number of Missed Trials* | 6.39<br>(1.03) | 4.99<br>(1.11) | 4.47<br>(1.03) | 6.31<br>(1.05) | Group: F(3, 192)=0.88, p=0.45, $\eta^2=0.01$<br>RTT MRT: F(1, 192)=9.41, **p=0.002**, $\eta^2=0.05$ |
| **RTT Mean Movement Time** | **EMM (SE)**<br>**n=49** | **EMM (SE)**<br>**n=48** | **EMM (SE)**<br>**n=50** | **EMM (SE)**<br>**n=50** | **ANCOVA** |
| *SST Number of Missed Trials* | 6.62<br>(1.07) | 6.14<br>(1.07) | 3.93<br>(1.06) | 5.51<br>(1.05) | Group: F(3, 192)=1.16, p=0.33, $\eta^2=0.02$<br>RTT MMT: F(1, 192)=0.08, p=0.78, $\eta^2=0.00$ |

**Key:** ANCOVA=Analysis of Covariance; ANOVA=Analysis of Variance; CC=clinical control; EMM=estimated marginal means; FMS=functional motor symptoms; FS=functional seizures; HC=healthy control; M=mean; SD=standard deviation; SE=standard error; WASI-II FSIQ-2=Wechsler Abbreviated Scale of Intelligence – Second edition Full Scale Intelligence Quotient – 2 subtest \*Welch's ANOVA

**Supplementary Table 20.**
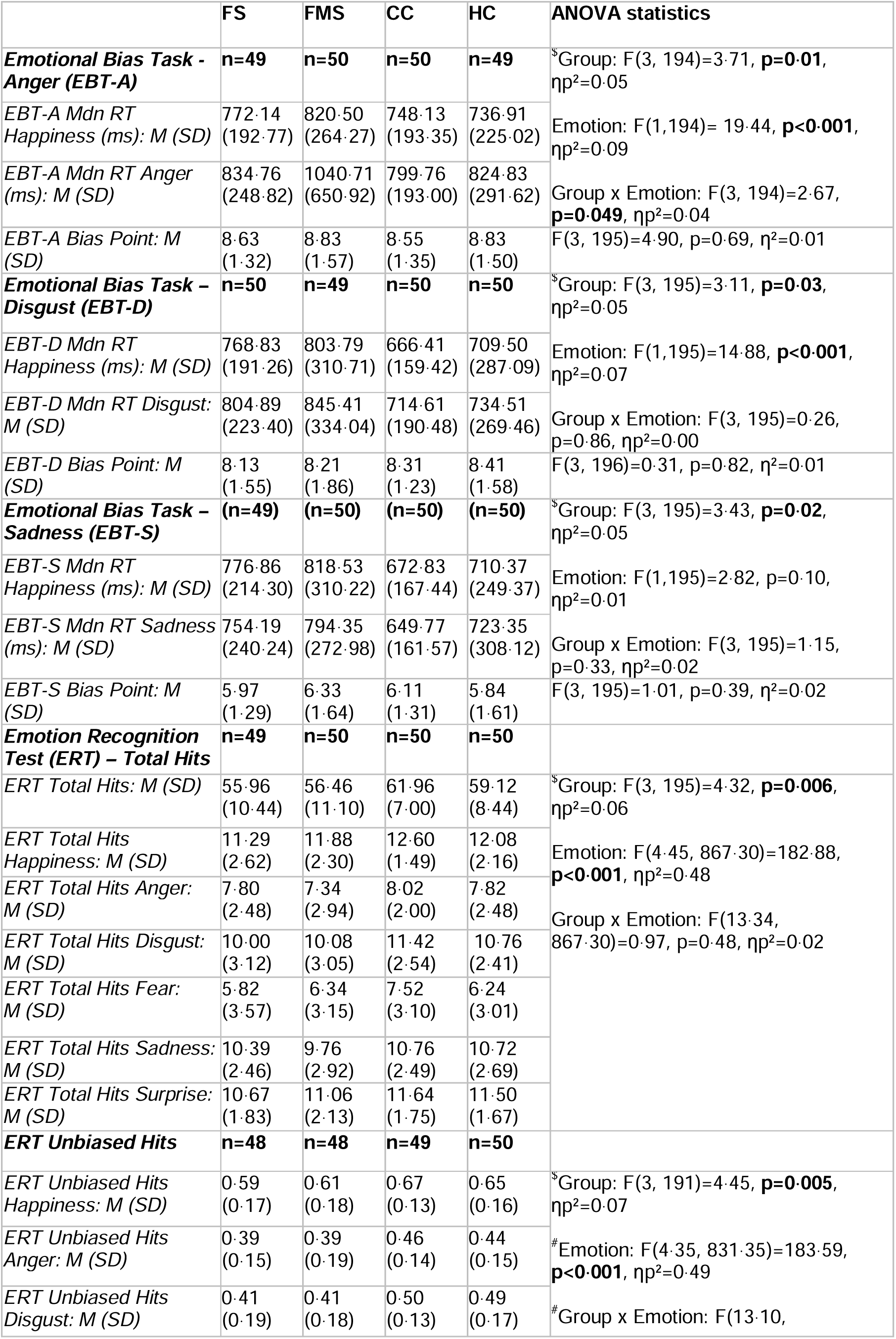
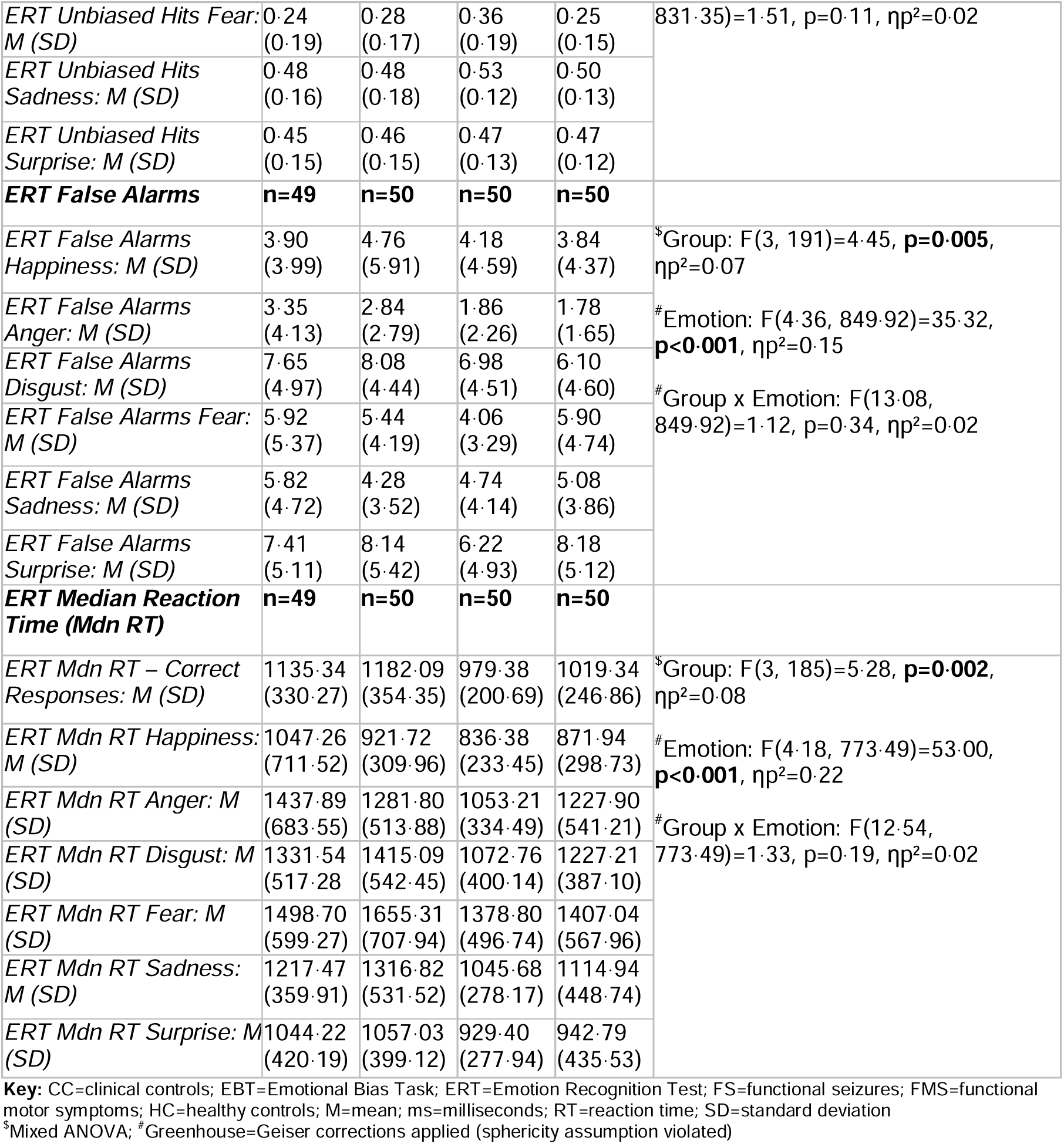
Emotional bias and recognition tasks uncorrected results.

**Supplementary Table 21.**
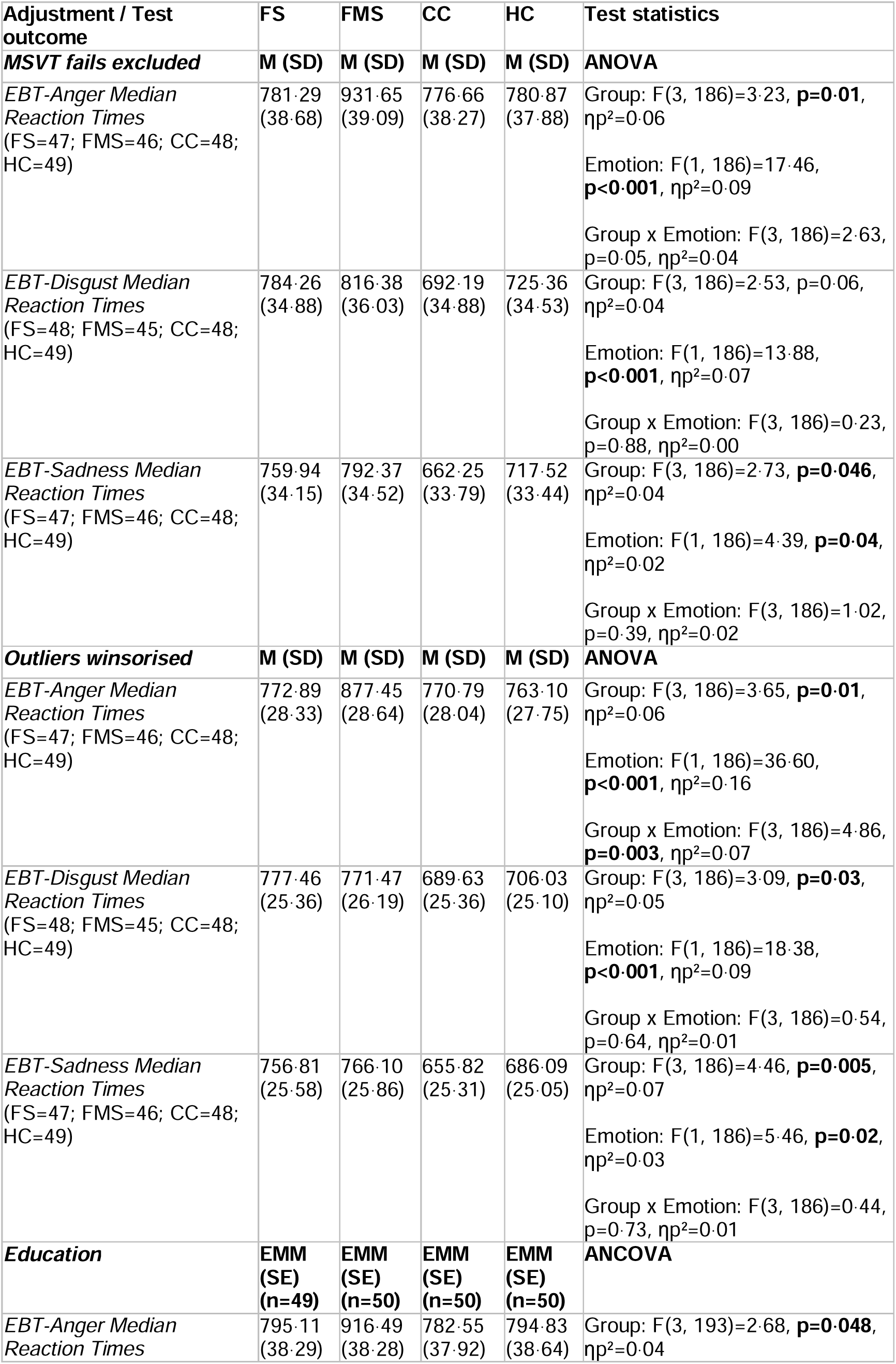
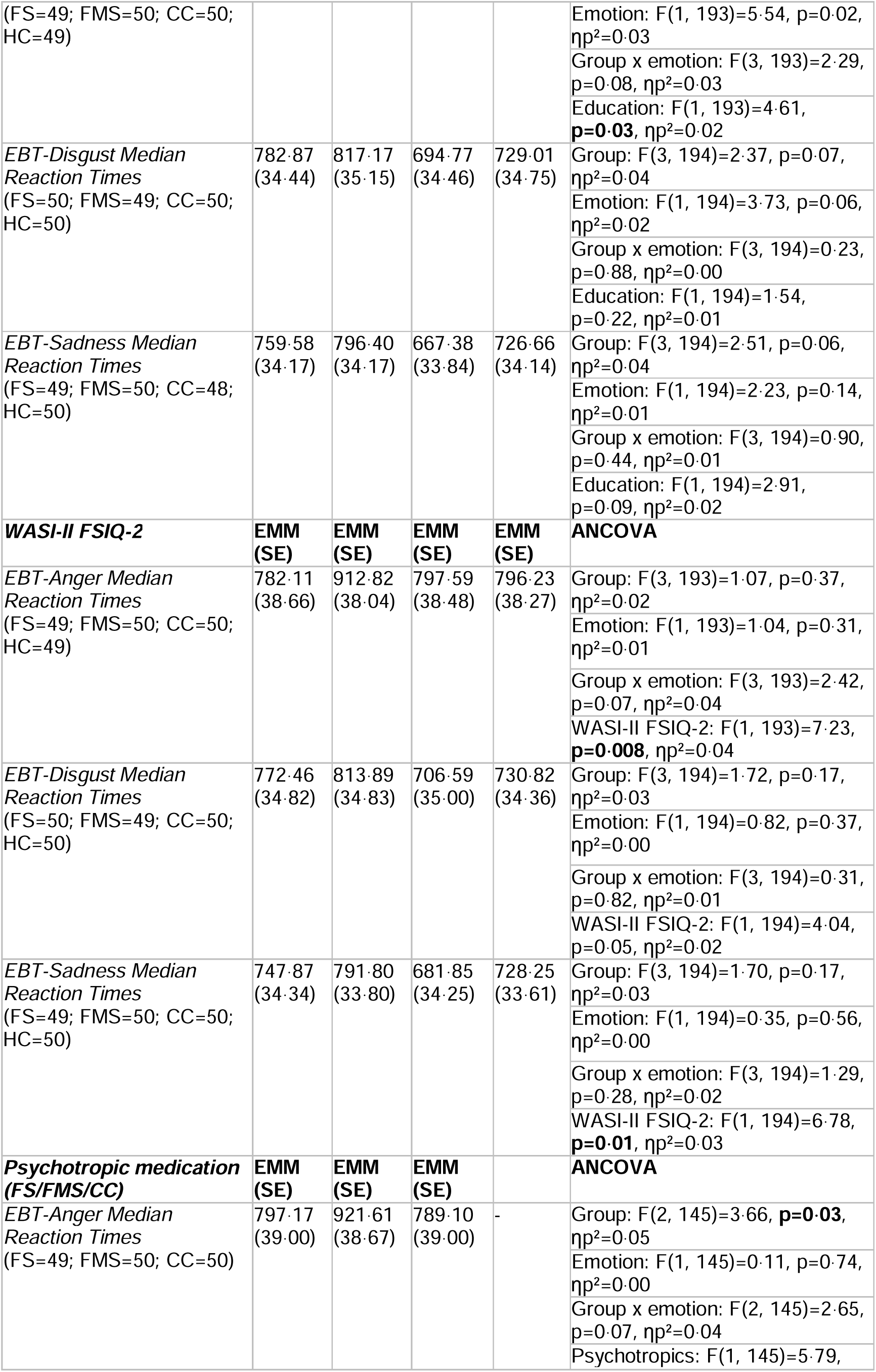
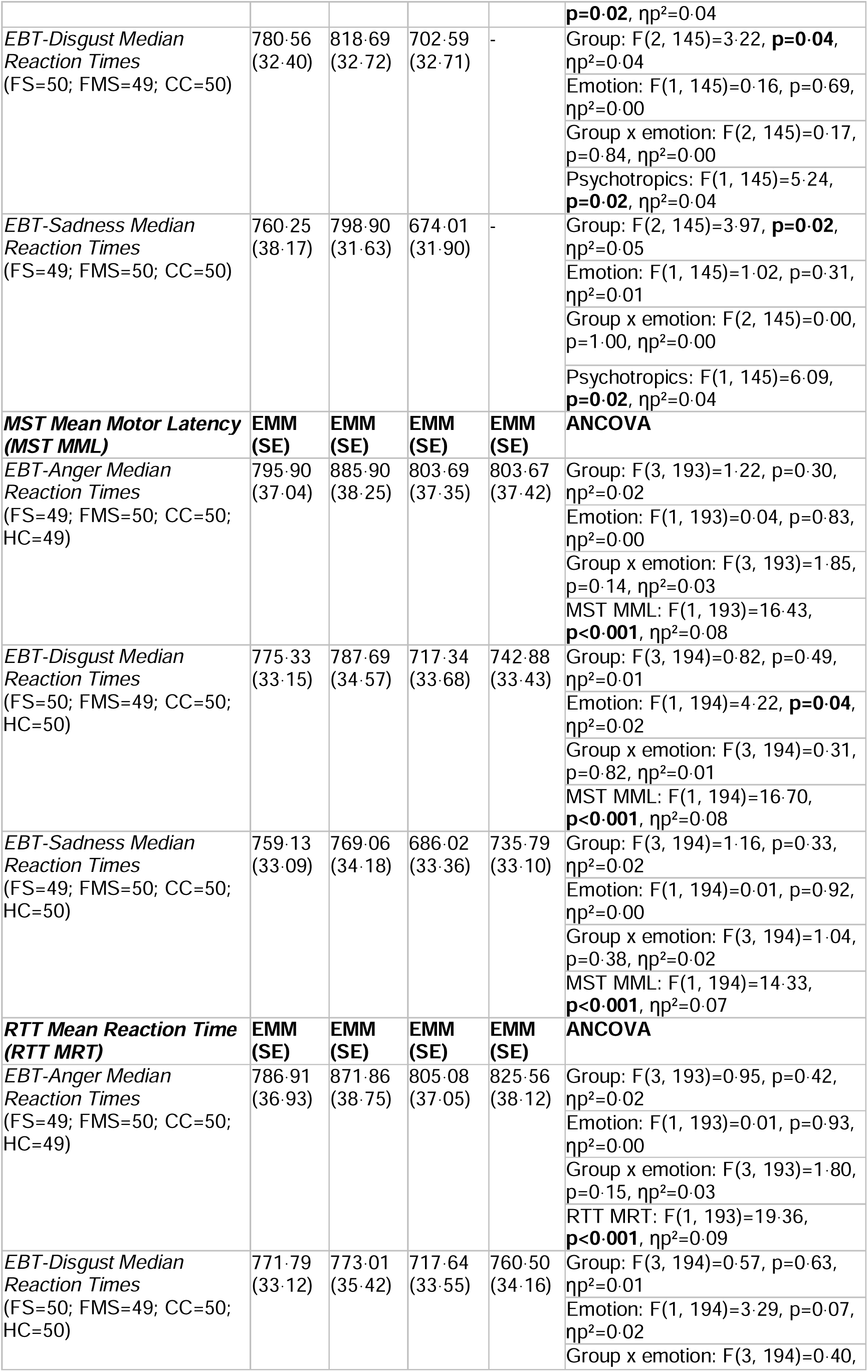
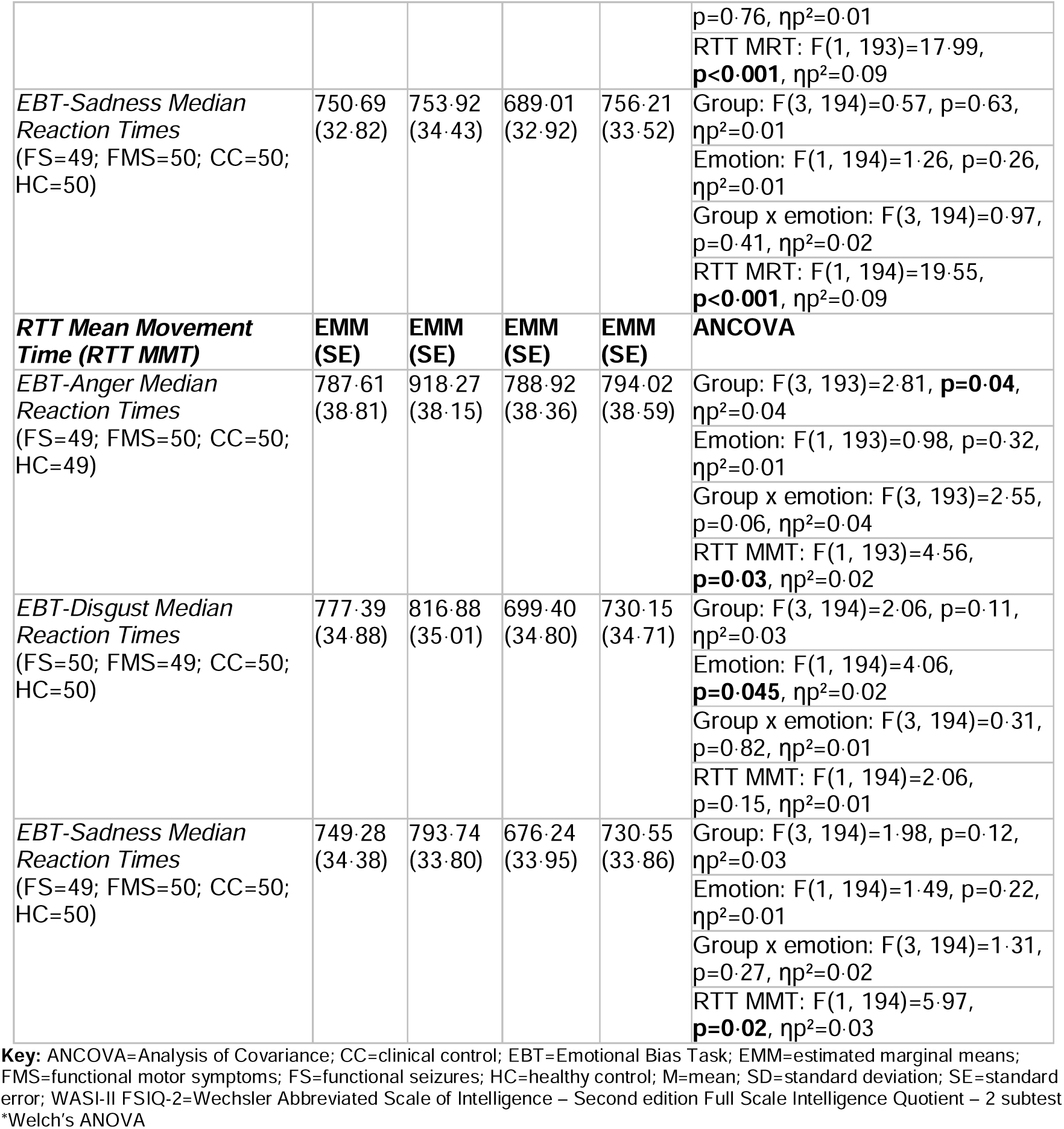
Emotional Bias Task sensitivity analyses.

**Supplementary Table 22.** Emotion Recognition Test sensitivity analyses.

| Adjustment / Test outcome | FS | FMS | CC | HC | Test statistics |
| --- | --- | --- | --- | --- | --- |
| **MSVT fails excluded** | **M (SD)** | **M (SD)** | **M (SD)** | **M (SD)** | **ANOVA** |
| *ERT Total Hits*<br>(FS=47; FMS=46;<br>CC=48; HC=49) | 9.51<br>(0.20) | 9.67<br>(0.20) | 10.37<br>(0.19) | 10.00<br>(0.19) | <sup>§</sup>Group: $F(3, 186)=3.86$, **$p=0.01$**,<br>$\eta^2=0.06$<br><sup>#</sup>Emotion: $F(4.38, 814.47)=175.35$,<br>**$p<0.001$**, $\eta^2=0.49$<br><sup>#</sup>Group x Emotion: $F(13.14, 814.47)=0.87$, $p=0.59$, $\eta^2=0.01$ |
| *ERT Unbiased Hits*<br>(FS=46; FMS=44;<br>CC=47; HC=49) | 0.44<br>(0.02) | 0.46<br>(0.02) | 0.51<br>(0.02) | 0.47<br>(0.02) | <sup>§</sup>Group: $F(3, 182)=3.94$, **$p=0.009$**,<br>$\eta^2=0.06$<br><sup>#</sup>Emotion: $F(4.37, 794.58)=176.98$,<br>**$p<0.001$**, $\eta^2=0.49$<br><sup>#</sup>Group x Emotion: $F(13.10, 794.58)=1.46$, $p=0.13$, $\eta^2=0.02$ |
| *ERT False Alarms*<br>(FS=47; FMS=46;<br>CC=48; HC=49) | 5.49<br>(0.20) | 5.33<br>(0.20) | 4.63<br>(0.19) | 5.00<br>(0.19) | <sup>§</sup>Group: $F(3, 186)=3.86$, **$p=0.01$**,<br>$\eta^2=0.06$<br><sup>#</sup>Emotion: $F(4.33, 806.02)=33.12$,<br>**$p<0.001$**, $\eta^2=0.15$<br><sup>#</sup>Group x Emotion: $F(13.00, 806.02)=1.08$, $p=0.37$, $\eta^2=0.02$ |
| *ERT Median Reaction Times*<br>FS=44; FMS=43;<br>CC=47; HC=47) | 1235.78<br>(45.64) | 1228.05<br>(46.17) | 1041.54<br>(44.16) | 1131.97<br>(44.16) | <sup>§</sup>Group: $F(3, 177)=4.17$, **$p=0.007$**,<br>$\eta^2=0.07$<br><sup>#</sup>Emotion: $F(3.94, 696.84)=64.55$,<br>**$p<0.001$**, $\eta^2=0.27$<br><sup>#</sup>Group x Emotion: $F(11.81, 696.84)=1.62$, $p=0.08$, $\eta^2=0.03$ |
| **Outliers winsorised** | **M (SD)** | **M (SD)** | **M (SD)** | **M (SD)** | **ANOVA** |
| *ERT Total Hits*<br>(FS=47; FMS=46;<br>CC=48; HC=49) | 9.52<br>(0.19) | 9.71<br>(0.19) | 10.38<br>(0.19) | 10.00<br>(0.19) | <sup>§</sup>Group: $F(3, 186)=3.88$, **$p=0.01$**,<br>$\eta^2=0.06$<br><sup>#</sup>Emotion: $F(4.26, 791.53)=188.13$,<br>**$p<0.001$**, $\eta^2=0.50$<br><sup>#</sup>Group x Emotion: $F(12.77, 791.53)=0.93$, $p=0.52$, $\eta^2=0.02$ |
| *ERT Unbiased Hits*<br>(FS=46; FMS=44;<br>CC=47; HC=49) | 0.44<br>(0.02) | 0.46<br>(0.02) | 0.47<br>(0.02) | 0.51<br>(0.02) | <sup>§</sup>Group: $F(3, 182)=4.09$, **$p=0.008$**,<br>$\eta^2=0.06$<br><sup>#</sup>Emotion: $F(4.35, 791.66)=181.58$,<br>**$p<0.001$**, $\eta^2=0.50$<br><sup>#</sup>Group x Emotion: $F(13.05, 791.66)=1.53$, $p=0.10$, $\eta^2=0.03$ |
| *ERT False Alarms*<br>(FS=47; FMS=46;<br>CC=48; HC=49) | 5.38<br>(0.18) | 5.24<br>(0.18) | 4.48<br>(0.18) | 4.85<br>(0.18) | <sup>§</sup> Group: $F(3, 186)=5.14$ , **$p=0.002$**,<br>$\eta^2=0.08$<br><sup>#</sup> Emotion: $F(4.24, 788.16)=40.95$ , **p<0.001**, $\eta^2=0.18$<br>#Group x Emotion: F(12.17, 788.16)=1.14, p=0.32, $\eta^2=0.02$ |
| *ERT Median Reaction Times*<br>(FS=44; FMS=43; CC=47; HC=47) | 1218.09<br>(39.50) | 1221.51<br>(39.95) | 1026.43<br>(38.21) | 1100.87<br>(38.21) | <sup>§</sup>Group: F(3, 177)=5.98, **p<0.001**, $\eta^2=0.09$<br>#Emotion: F(3.55, 627.45)=84.86, **p<0.001**, $\eta^2=0.32$<br>#Group x Emotion: F(10.64, 627.45)=2.14, **p=0.02**, $\eta^2=0.04$ |
| **Education** | **EMM (SE)** | **EMM (SE)** | **EMM (SE)** | **EMM (SE)** | **ANCOVA** |
| *ERT Total Hits*<br>(FS=49; FMS=50; CC=50; HC=50) | 9.35<br>(0.23) | 9.45<br>(0.22) | 10.30<br>(0.22) | 9.82<br>(0.22) | Group: F(3, 194)=3.63, **p=0.01**, $\eta^2=0.05$<br>#Emotion: F(4.45, 862.52)=21.04, **p<0.001**, $\eta^2=0.10$<br>#Group x emotion: F(13.34, 862.52)=1.00, p=0.45, $\eta^2=0.02$<br>Education: F(1, 194)=0.98, p=0.32, $\eta^2=0.01$ |
| *ERT Unbiased Hits*<br>(FS=48; FMS=48; CC=49; HC=50) | 0.43<br>(0.02) | 0.44<br>(0.02) | 0.51<br>(0.02) | 0.46<br>(0.02) | Group: F(3, 190)=3.80, **p=0.01**, $\eta^2=0.06$<br>#Emotion: F(4.35, 826.39)=23.08, **p<0.001**, $\eta^2=0.11$<br>#Group x emotion: F(13.05, 826.39)=1.48, p=0.12, $\eta^2=0.02$<br>Education: F(1, 190)=0.88, p=0.35, $\eta^2=0.01$ |
| *ERT False Alarms*<br>(FS=49; FMS=50; CC=50; HC=50) | 5.65<br>(0.23) | 5.55<br>(0.22) | 4.70<br>(0.22) | 5.18<br>(0.22) | Group: F(3, 194)=3.63, **p=0.01**, $\eta^2=0.05$<br>#Emotion: F(4.36, 844.93)=4.00, **p=0.002**, $\eta^2=0.02$<br>#Group x emotion: F(13.07, 844.93)=1.08, p=0.37, $\eta^2=0.02$<br>Education: F(1, 194)=0.98, p=0.32, $\eta^2=0.01$ |
| *ERT Median Reaction Times*<br>(FS=46; FMS=47; CC=49; HC=47) | 1254.01<br>(47.33) | 1258.27<br>(47.41) | 1062.04<br>(45.91) | 1147.23<br>(47.31) | Group: F(3, 184)=3.92, **p=0.01**, $\eta^2=0.06$<br>#Emotion: F(4.18, 769.70)=3.92, **p=0.003**, $\eta^2=0.02$<br>#Group x emotion: F(12.55, 769.70)=1.34, p=0.19, $\eta^2=0.02$<br>Education: F(1, 184)=3.46, **p=0.06**, $\eta^2=0.02$ |
| **WASI-II FSIQ-2** | **EMM (SE)** | **EMM (SE)** | **EMM (SE)** | **EMM (SE)** | **ANCOVA** |
| *ERT Total Hits*<br>(FS=49; FMS=50; CC=50; HC=50) | 9.54<br>(0.22) | 9.59<br>(0.21) | 10.08<br>(0.21) | 9.71<br>(0.21) | Group: F(3, 194)=1.21, p=0.31, $\eta^2=0.02$<br>#Emotion: F(4.46, 864.59)=5.18, **p<0.001**, $\eta^2=0.03$<br>#Group x emotion: F(13.37, 864.59)=0.84, p=0.63, $\eta^2=0.01$<br>WASI-II FSIQ-2: F(1, 194)=25.91, **p<0001**, $\eta^2=0.12$ |
| *ERT Unbiased Hits*<br>(FS=48; FMS=48;<br>CC=49; HC=50) | 0.44<br>(0.02) | 0.45<br>(0.02) | 0.49<br>(0.02) | 0.46<br>(0.02) | Group: F(3, 190)=1.37, p=0.26, $\eta^2=0.02$<br>#Emotion: F(4.34, 824.14)=7.02, **p<0.001**, $\eta^2=0.04$<br>#Group x emotion: F(13.01, 824.14)=1.35, p=0.18, $\eta^2=0.02$<br>WASI-II FSIQ-2: F(1, 190)=24.56, **p<0.001**, $\eta^2=0.11$ |
| *ERT False Alarms*<br>(FS=49; FMS=50;<br>CC=50; HC=50) | 5.46<br>(0.22) | 5.41<br>(0.21) | 4.92<br>(0.21) | 5.29<br>(0.21) | Group: F(3, 194)=1.21, p=0.31, $\eta^2=0.02$<br>#Emotion: F(4.37, 847.13)=3.13, **p=0.01**, $\eta^2=0.02$<br>#Group x emotion: F(13.10, 847.13)=1.05, p=0.40, $\eta^2=0.02$<br>WASI-II FSIQ-2: F(1, 194)=25.91, **p<0.001**, $\eta^2=0.12$ |
| *ERT Median Reaction Times*<br>FS=44; FMS=43;<br>CC=47; HC=47) | 1235.46<br>(47.22) | 1244.87<br>(46.89) | 1084.49<br>(46.11) | 1155.40<br>(46.50) | Group: F(3, 184)=2.39, p=0.07, $\eta^2=0.04$<br>#Emotion: F(4.17, 767.55)=1.81, p=0.12, $\eta^2=0.01$<br>#Group x emotion: F(12.51, 767.55)=1.25, p=0.24, $\eta^2=0.02$<br>WASI-II FSIQ-2: F(1, 184)=9.40, **p=0.003**, $\eta^2=0.04$ |
| **Psychotropic medications (FS/FMS/CC)** | **EMM (SE)** | **EMM (SE)** | **EMM (SE)** |  | **ANCOVA** |
| *ERT Total Hits*<br>(FS=49; FMS=50;<br>CC=50) | 9.35<br>(0.23) | 9.44<br>(0.23) | 10.28<br>(0.23) | - | Group: F(2, 145)=4.85, **p=0.009**, $\eta^2=0.06$<br>#Emotion: F(4.35, 631.30)=15.74, **p<0.001**, $\eta^2=0.10$<br>#Group x emotion: F(8.71, 631.30)=1.07, p=0.38, $\eta^2=0.02$<br>Psychotropics: F(1, 145)=1.81, p=0.18, $\eta^2=0.01$ |
| *ERT Unbiased Hits*<br>(FS=48; FMS=48;<br>CC=49) | 0.43<br>(0.02) | 0.44<br>(0.02) | 0.50<br>(0.02) | - | Group: F(2, 141)=5.27, **p=0.006**, $\eta^2=0.07$<br>#Emotion: F(4.38, 617.41)=17.44, **p<0.001**, $\eta^2=0.11$<br>#Group x emotion: F(8.76, 617.41)=0.52, p=0.85, $\eta^2=0.01$<br>Psychotropics: F(1, 141)=1.15, p=0.29, $\eta^2=0.01$ |
| *ERT False Alarms*<br>(FS=49; FMS=50;<br>CC=50) | 5.65<br>(0.23) | 5.56<br>(0.23) | 4.72<br>(0.23) | - | Group: F(2, 145)=4.85, **p=0.009**, $\eta^2=0.06$<br>#Emotion: F(4.43, 642.32)=2.87, **p=0.02**, $\eta^2=0.02$<br>#Group x emotion: F(8.86, 642.32)=0.75, p=0.66, $\eta^2=0.01$<br>Psychotropics: F(1, 144)=1.81, p=0.18, $\eta^2=0.01$ |
| *ERT Median Reaction Times*<br>FS=46; FMS=47;<br>CC=49) | 1254.70<br>(48.02) | 1268.19<br>(47.50) | 1066.53<br>(46.98) | - | Group: F(2, 138)=5.59, **p=0.005**, $\eta^2=0.08$<br>#Emotion: F(3.96, 545.93)=8.40, **p<0.001**, $\eta^2=0.06$<br>#Group x emotion: F(7.91, 545.93)=1.82, p=0.07, $\eta^2=0.03$<br>Psychotropics: F(1, 138)=3.47, p=0.07, $\eta p^2=0.03$ |
| **MST Mean Motor Latency (MST MML)** | **EMM (SE)** | **EMM (SE)** | **EMM (SE)** | **EMM (SE)** | **ANCOVA** |
| ERT Total Hits<br>(FS=49; FMS=50;<br>CC=50; HC=50) | 9.36<br>(0.22) | 9.61<br>(0.23) | 10.20<br>(0.22) | 9.75<br>(0.22) | Group: F(3, 194)=2.47, p=0.06,<br>$\eta p^2=0.04$<br>#Emotion: F(4.45, 862.24)=15.80,<br>**p<0.001**, $\eta p^2=0.08$<br>#Group x emotion: F(13.33,<br>862.24)=0.99, p=0.46, $\eta p^2=0.02$<br>MST MML: F(1, 194)=8.90, **p=0.003**,<br>$\eta p^2=0.04$ |
| ERT Unbiased Hits<br>(FS=48; FMS=48;<br>CC=49; HC=50) | 0.43<br>(0.02) | 0.45<br>(0.02) | 0.50<br>(0.02) | 0.46<br>(0.02) | Group: F(3, 190)=2.76, **p=0.04**,<br>$\eta p^2=0.04$<br>#Emotion: F(4.35, 826.58)=19.66,<br>**p<0.001**, $\eta p^2=0.09$<br>#Group x emotion: F(13.05,<br>826.58)=1.42, p=0.15, $\eta p^2=0.02$<br>MST MML: F(1, 190)=7.67, **p=0.006**,<br>$\eta p^2=0.04$ |
| ERT False Alarms<br>(FS=49; FMS=50;<br>CC=50; HC=50) | 5.64<br>(0.22) | 5.40<br>(0.23) | 4.80<br>(0.22) | 5.25<br>(0.22) | Group: F(3, 194)=2.47, p=0.06,<br>$\eta p^2=0.04$<br>#Emotion: F(4.36, 844.86)=3.31,<br>**p=0.009**, $\eta p^2=0.02$<br>#Group x emotion: F(13.07,<br>844.86)=1.14, p=0.32, $\eta p^2=0.02$<br>MST MML: F(1, 194)=8.90, **p=0.003**,<br>$\eta p^2=0.04$ |
| ERT Median Reaction Times<br>FS=46; FMS=47;<br>CC=49; HC=47) | 1249.33<br>(45.27) | 1219.57<br>(46.37) | 1092.17<br>(44.65) | 1159.11<br>(45.10) | Group: F(3, 184)=2.27, p=0.08,<br>$\eta p^2=0.04$<br>#Emotion: F(4.17, 766.75)=5.66,<br>**p<0.001**, $\eta p^2=0.03$<br>#Group x emotion: F(12.50,<br>766.75)=1.41, p=0.15, $\eta p^2=0.02$<br>MST MML: F(1, 184)=19.79, **p<0.001**,<br>$\eta p^2=0.10$ |
| **RTT Mean Reaction Time (RTT MRT)** | **EMM (SE)** | **EMM (SE)** | **EMM (SE)** | **EMM (SE)** | **ANCOVA** |
| ERT Total Hits<br>(FS=49; FMS=50;<br>CC=50; HC=50) | 9.47<br>(0.20) | 9.90<br>(0.21) | 10.07<br>(0.20) | 9.48<br>(0.21) | Group: F(3, 194)=2.26, p=0.08,<br>$\eta p^2=0.03$<br>#Emotion: F(4.45, 863.71)=4.95,<br>**p<0.001**, $\eta p^2=0.03$<br>#Group x emotion: F(13.36,<br>863.71)=1.02, p=0.43, $\eta p^2=0.02$<br>RTT MRT: F(1, 194)=45.08, **p<0.001**,<br>$\eta p^2=0.19$ |
| ERT Unbiased Hits<br>(FS=48; FMS=48;<br>CC=49; HC=50) | 0.44<br>(0.02) | 0.47<br>(0.02) | 0.49<br>(0.02) | 0.44<br>(0.02) | Group: F(3, 190)=2.77, **p=0.04**,<br>$\eta p^2=0.04$<br>#Emotion: F(4.36, 828.27)=6.64,<br>**p<0.001**, $\eta p^2=0.03$<br>#Group x emotion: F(13.08,<br>828.27)=1.39, p=0.16, $\eta p^2=0.02$<br>RTT MRT: F(1, 190)=41.90, **p<0.001**,<br>$\eta p^2=0.18$ |
| ERT False Alarms<br>(FS=49; FMS=50;<br>CC=50; HC=50) | 5.53<br>(0.20) | 5.10<br>(0.21) | 4.93<br>(0.20) | 5.52<br>(0.21) | Group: F(3, 194)=2.26, p=0.08,<br>$\eta p^2=0.03$<br>#Emotion: F(4.35, 844.19)=1.68,<br>p=0.15, $\eta p^2=0.01$<br>#Group x emotion: F(13.05, |
| | | | | | 844.19)=1.27, $p=0.23$, $\eta^2=0.02$<br>RTT MRT: $F(1, 194)=45.08$, **$p<0.001$**, $\eta^2=0.19$ |
| *ERT Median Reaction Times*<br>FS=46; FMS=47;<br>CC=49; HC=47) | 1236.27<br>(44.07) | 1183.22<br>(46.23) | 1099.41<br>(43.25) | 1200.69<br>(45.00) | Group: $F(3, 184)=1.79$, $p=0.15$, $\eta^2=0.03$<br>#Emotion: $F(4.22, 777.11)=10.85$, **$p<0.001$**, $\eta^2=0.06$<br>#Group x emotion: $F(12.67, 777.11)=1.97$, **$p=0.02$**, $\eta^2=0.03$<br>RTT MRT: $F(1, 184)=32.48$, **$p<0.001$**, $\eta^2=0.15$ |
| **RTT Mean Movement Time (RTT MMT)** | **EMM (SE)** | **EMM (SE)** | **EMM (SE)** | **EMM (SE)** | **ANCOVA** |
| *ERT Total Hits*<br>(FS=49; FMS=50;<br>CC=50; HC=50) | 9.43<br>(0.23) | 9.49<br>(0.22) | 10.23<br>(0.22) | 9.77<br>(0.22) | Group: $F(3, 194)=2.55$, $p=0.06$, $\eta^2=0.04$<br>#Emotion: $F(4.45, 862.93)=9.95$, **$p<0.001$**, $\eta^2=0.05$<br>#Group x emotion: $F(13.34, 862.93)=0.92$, $p=0.54$, $\eta^2=0.01$<br>RTT MMT: $F(1, 194)=5.59$, **$p=0.02$**, $\eta^2=0.03$ |
| *ERT Unbiased Hits*<br>(FS=48; FMS=48;<br>CC=49; HC=50) | 0.44<br>(0.02) | 0.45<br>(0.02) | 0.50<br>(0.02) | 0.46<br>(0.02) | Group: $F(3, 190)=2.47$, $p=0.06$, $\eta^2=0.04$<br>#Emotion: $F(4.36, 828.99)=5.98$, **$p<0.001$**, $\eta^2=0.03$<br>#Group x emotion: $F(13.09, 828.99)=1.53$, $p=1.00$, $\eta^2=0.02$<br>RTT MMT: $F(1, 190)=9.04$, **$p=0.003$**, $\eta^2=0.05$ |
| *ERT False Alarms*<br>(FS=49; FMS=50;<br>CC=50; HC=50) | 5.57<br>(0.23) | 5.51<br>(0.22) | 4.77<br>(0.22) | 5.23<br>(0.22) | Group: $F(3, 194)=2.55$, $p=0.06$, $\eta^2=0.04$<br>#Emotion: $F(4.36, 845.60)=3.57$, **$p=0.005$**, $\eta^2=0.02$<br>#Group x emotion: $F(13.08, 845.60)=1.17$, $p=0.29$, $\eta^2=0.02$<br>RTT MMT: $F(1, 194)=5.59$, **$p=0.02$**, $\eta^2=0.03$ |
| *ERT Median Reaction Times*<br>FS=46; FMS=47;<br>CC=49; HC=47) | 1237.60<br>(46.63) | 1241.68<br>(46.54) | 1083.74<br>(45.51) | 1157.27<br>(46.14) | Group: $F(3, 184)=2.48$, $p=0.06$, $\eta^2=0.04$<br>#Emotion: $F(4.17, 768.00)=4.11$, **$p=0.002$**, $\eta^2=0.02$<br>#Group x emotion: $F(12.52, 768.00)=1.23$, $p=0.26$, $\eta^2=0.02$<br>RTT MMT: $F(1, 184)=12.04$, **$p<0.001$**, $\eta^2=0.06$ |

**Key:** ANCOVA=Analysis of Covariance; CC=clinical control; ERT=Emotion Recognition Test; EMM=estimated marginal means; FMS=functional motor symptoms; FS=functional seizures; HC=healthy control; SE=standard error; WASI-II FSIQ-2=Wechsler Abbreviated Scale of Intelligence – Second edition Full Scale Intelligence Quotient – 2 subtest
<sup>s</sup>Mixed ANOVA; #Greenhouse-Geiser correction (sphericity assumption violated)

**Supplementary Table 23.** Metacognitive performance ratings.

| Test | FS<br>(n=50) | FMS<br>(n=50) | CC<br>(n=50) | HC<br>(n=50) | ANOVA statistics |
| --- | --- | --- | --- | --- | --- |
| **Medical Symptom Validity Test: M (SD)** | 5.13<br>(1.08) | 5.16<br>(1.13) | 5.04<br>(1.09) | 5.18<br>(1.04) | F(3, 194)=0.16, p=0.92, $\eta^2=0.00$ |
| **WASI-II Vocabulary: M (SD)** | 4.48<br>(0.79) | 4.34<br>(0.98) | 4.80<br>(0.93) | 4.50<br>(0.86) | F(3, 196)=2.35, p=0.07, $\eta^2=0.04$ |
| **WASI-II Matrix Reasoning: M (SD)** | 3.74<br>(1.14) | 3.62<br>(1.16) | 3.92<br>(1.01) | 4.14<br>(0.99) | F(3, 196)=2.21, p=0.09, $\eta^2=0.03$ |
| **Motor Screening Test: M (SD)** | 5.52<br>(1.11) | 5.62<br>(1.21) | 5.12<br>(1.27) | 5.32<br>(1.19) | F(3, 196)=1.72, p=0.17, $\omega^2=0.03$ |
| **Reaction Time Test: M (SD) FS=49<sup>§</sup>** | 4.74<br>(1.17) | 4.80<br>(1.40) | 4.72<br>(1.14) | 5.12<br>(1.12) | F(3, 195)=1.19, p=0.31, $\eta^2=0.02$ |
| **Rapid Visual Information Processing (RVIP): M (SD) FS=47; CC/HC=49<sup>§</sup>** | 2.94<br>(1.05) | 2.40<br>(1.14) | 3.53<br>(1.23) | 3.88<br>(1.18) | F(3, 192)=15.79, **p<0.001**, $\eta^2=0.20$ |
| **Spatial Span Forward: M (SD) HC=49<sup>§</sup>** | 3.18<br>(1.08) | 3.36<br>(1.22) | 3.36<br>(1.08) | 3.57<br>(1.19) | F(3, 195)=0.96, p=0.41, $\eta^2=0.02$ |
| **Spatial Span Reverse: M (SD)** | 3.36<br>(1.34) | 2.96<br>(1.31) | 3.26<br>(1.17) | 3.54<br>(1.13) | F(3, 196)=1.92, p=0.13, $\eta^2=0.03$ |
| **Intra-Extra Dimensional Set Shift (IEDSS): M (SD)** | 3.92<br>(1.58) | 4.04<br>(1.36) | 4.56<br>(0.91) | 4.44<br>(1.11) | F(3, 196)=2.98, p<0.03<sup>†</sup>, $\eta^2=0.04$ |
| **Stop Signal Task (SST): M (SD) FS/FMS=49<sup>§</sup>** | 3.53<br>(1.06) | 3.50<br>(1.14) | 3.48<br>(1.00) | 3.94<br>(1.06) | F(3, 194)=2.15, p=0.10, $\eta^2=0.03$ |
| **Emotional Bias Task – Anger: M (SD) FS=49<sup>§</sup>** | 4.67<br>(1.13) | 4.48<br>(1.33) | 4.56<br>(0.95) | 4.46<br>(0.86) | *F(3, 107.07)=0.41, p=0.75, $\omega^2=0.01$ |
| **Emotional Bias Task – Disgust: M (SD) FS=49<sup>§</sup>** | 4.18<br>(1.17) | 4.34<br>(1.24) | 4.52<br>(0.71) | 4.34<br>(0.92) | *F(3, 105.58)=1.14, p=0.34, $\omega^2=0.00$ |
| **Emotional Bias Task – Sadness: M (SD) FS=49<sup>§</sup>** | 4.18<br>(1.01) | 4.46<br>(1.15) | 4.46<br>(0.76) | 4.44<br>(0.94) | F(3, 195)=0.99, p=0.40, $\eta^2=0.02$ |
| **Emotion Recognition Test: M (SD) FS=49<sup>§</sup>** | 3.53<br>(1.12) | 3.62<br>(1.10) | 3.88<br>(0.80) | 3.90<br>(1.04) | *F(3, 107.10)=1.61, p=0.19, $\omega^2=0.01$ |

**Key:** CC=clinical controls; FS=functional seizures; FMS=functional motor symptoms; HC=healthy controls; M=mean; SD=standard deviation <sup>§</sup>Sample size deviated due to missing data \*Welch's ANOVA <sup>†</sup>Did not survive Benjamini-Hochberg correction (5%)

**Supplementary Table 24.** RVIP performance ratings sensitivity analyses.

| Adjustment | FS | FMS | CC | HC | Test statistics |
| --- | --- | --- | --- | --- | --- |
| **MSVT fails excluded** | **M (SD)**<br>**n=46** | **M (SD)**<br>**n=46** | **M (SD)**<br>**n=47** | **M (SD)**<br>**n=48** | **ANOVA** |
| | 2.98<br>(1.02) | 2.46<br>(1.15) | 3.60<br>(1.20) | 3.88<br>(1.20) | F(3, 183)=14.47, **p<0.001**, $\eta^2=0.19$ |
| **Outliers winsorised** | **M (SD)**<br>**n=46** | **M (SD)**<br>**n=46** | **M (SD)**<br>**n=47** | **M (SD)**<br>**n=48** | **ANOVA** |
| | 3.00<br>(1.02) | 2.46<br>(1.15) | 3.57<br>(1.14) | 3.813<br>(1.10) | F(3, 183)=14.28, **p<0.001**, $\eta^2=0.19$ |
| **Education** | **EMM**<br>**(SE)**<br>**n=47** | **EMM**<br>**(SE)**<br>**n=50** | **EMM**<br>**(SE)**<br>**n=49** | **EMM**<br>**(SE)**<br>**n=49** | **ANCOVA** |
| | 2.97<br>(0.17) | 2.46<br>(0.17) | 3.49<br>(0.17) | 3.82<br>(0.17) | Group: F(3, 190)=12.43, **p<0.001**, $\eta^2=0.16$<br>Education: F(1, 190)=4.36, **p=0.04**, $\eta^2=0.02$ |
| **WASI-II FSIQ-2** | **EMM**<br>**(SE)**<br>**n=47** | **EMM**<br>**(SE)**<br>**n=50** | **EMM**<br>**(SE)**<br>**n=49** | **EMM**<br>**(SE)**<br>**n=49** | **ANCOVA** |
| | 3.04<br>(0.17) | 2.49<br>(0.16) | 3.41<br>(0.17) | 3.81<br>(0.16) | Group: F(3, 190)=11.42, **p<0.001**, $\eta^2=0.15$<br>WASI-II FSIQ-2: F(1, 190)=9.40, **p=0.002**, $\eta^2=0.05$ |
| **Psychotropic medication** | **EMM**<br>**(SE)**<br>**n=47** | **EMM**<br>**(SE)**<br>**n=50** | **EMM**<br>**(SE)**<br>**n=49** |  | **ANCOVA** |
| | 2.96<br>(0.17) | 2.43<br>(0.16) | 3.47<br>(0.16) | - | Group: F(2, 142)=10.19, **p<0.001**, $\eta^2=0.13$<br>Psychotropics: F(1, 142)=5.54, **p=0.02**, $\eta^2=0.04$ |
| **RVIP Ability** | **EMM**<br>**(SE)**<br>**n=49** | **EMM**<br>**(SE)**<br>**n=48** | **EMM**<br>**(SE)**<br>**n=50** | **EMM**<br>**(SE)**<br>**n=50** |  |
| | 3.11<br>(0.16) | 2.69<br>(0.16) | 3.37<br>(0.16) | 3.75<br>(0.16) | Group: F(3, 182)=7.35, **p<0.001**, $\eta^2=0.11$<br>RVIP Ability: F(1, 182)=30.34, **p<0.001**, $\eta^2=0.14$ |

**Key:** ANCOVA=Analysis of Covariance; CC=clinical control; EMM=estimated marginal means; FMS=functional motor symptoms; FS=functional seizures; HC=healthy control; M=mean; RVIP=Rapid Visual Information Processing; SD=standard deviation; SE=standard error; WASI-II FSIQ-2=Wechsler Abbreviated Scale of Intelligence – Second edition Full Scale Intelligence Quotient – 2 subtest

**Supplementary Table 25.** RVIP performance ratings post-hoc tests.

| Comparison | Mean difference (standard error) | Confidence interval (95%) | p-value |
| --- | --- | --- | --- |
| FS vs FMS | 0.54 (0.23) | -0.09, 1.16 | 0.14 |
| FS vs HC | -0.94 (0.24) | -1.57, -0.31 | **<0.001** |
| FS vs CC | -0.59 (0.24) | -1.22, 0.03 | 0.08 |
| FMS vs HC | -1.48 (0.23) | -2.10, -0.86 | **<0.001** |
| FMS vs CC | -1.13 (0.23) | -1.75, -0.51 | **<0.001** |
| HC vs CC | 0.35 (0.23) | -0.27, 0.97 | 0.83 |

**Key:** CC=clinical controls; FMS=functional motor symptoms; FS=functional seizures; HC=healthy controls

**Supplementary Table 26.** Significant within-group correlations.

| Outcome variable | FS |  | FMS |  | CC |  |
| --- | --- | --- | --- | --- | --- | --- |
|  | *r/r<sub>s</sub>* | *p* | *r/r<sub>s</sub>* | *p* | *r/r<sub>s</sub>* | *p* |
| **CFQ Total** |  |  |  |  |  |  |
| GAD-7 | 0.38 | **0.008** | 0.38 | **0.006** | 0.34 | **0.02** |
| PHQ-9 | 0.45 | **0.001** | 0.48 | **<0.001** | 0.45 | **0.001** |
| MDI Disengagement *T* | 0.59 | **<0.001** | 0.63 | **<0.001** | 0.49 | **<0.001** |
| MDI Depersonalisation *T* | 0.42 | **0.003** | 0.34 | **0.02** | 0.28 | 0.06 |
| MDI Derealisation *T* | 0.42 | **0.003** | 0.41 | **0.003** | 0.21 | 0.16 |
| MDI Emotional Constriction *T* | 0.32 | 0.03<sup>†</sup> | 0.29 | 0.04<sup>†</sup> | 0.37 | **0.01** |
| MDI Memory Disturbance *T* | 0.46 | **<0.001** | 0.59 | **<0.001** | 0.36 | **0.01** |
| MDI Identity Disturbance *T* | 0.22 | 0.12 | 0.46 | **<0.001** | 0.18 | 0.23 |
| SDQ-20 | 0.45 | **0.001** | 0.46 | **<0.001** | 0.43 | **0.002** |
| PHQ-15 | 0.45 | **0.001** | 0.23 | 0.11 | 0.39 | **0.006** |
| FNSQ Total | 0.02 | 0.88 | 0.43 | **0.002** | NA | NA |
| FNSQ Severity | -0.09 | 0.53 | 0.34 | **0.02** | NA | NA |
| FNSQ Impact | -0.20 | 0.17 | 0.30 | **0.03** | NA | NA |
| WSAS | -0.03 | 0.83 | 0.39 | **0.005** | 0.50 | **<0.001** |
| SF-36 Physical Function | -0.07 | 0.66 | -0.38 | **0.007** | -0.57 | **<0.001** |
| SF-36 Role Limitations – Physical | -0.17 | 0.26 | -0.50 | **<0.001** | -0.43 | **0.003** |
| SF-36 Role Limitations – Emotional | -0.43 | **0.002** | -0.44 | **0.002** | -0.48 | **<0.001** |
| SF-36 Energy | -0.22 | 0.15 | -0.33 | **0.03** | -0.27 | 0.06 |
| SF-36 Emotional Wellbeing | -0.35 | **0.02** | -0.43 | **0.002** | -0.34 | **0.02** |
| SF-36 Social Functioning | -0.11 | 0.45 | -0.47 | **<0.001** | -0.42 | 0.003 |
| SF-36 Pain | -0.18 | 0.23 | -0.48 | **<0.001** | -0.52 | **<0.001** |
| SF-36 General Health | -0.53 | **<0.001** | -0.36 | **0.01** | -0.41 | **0.004** |
| **RVIP Performance ratings** |  |  |  |  |  |  |
| CFQ Total | -0.09 | 0.55 | -0.43 | **0.002** | -0.22 | 0.14 |
| RVIP Ability | 0.27 | 0.07 | 0.48 | **<0.001** | 0.24 | 0.10 |
| RVIP Probability of Hit | 0.19 | 0.21 | 0.36 | **0.01** | 0.22 | 0.12 |
| RVIP Total Misses | -0.19 | 0.21 | -0.36 | **0.01** | -0.22 | 0.12 |
| PHQ-9 | 0.14 | 0.37 | -0.39 | **0.005** | -0.21 | 0.15 |
| GAD-7 | 0.02 | 0.88 | -0.37 | **0.007** | -0.09 | 0.53 |
| MDI Disengagement *T* | -0.10 | 0.51 | -0.53 | **<0.001** | -0.11 | 0.45 |
| MDI Depersonalisation *T* | 0.17 | 0.25 | -0.36 | **0.01** | -0.00 | 0.98 |
| MDI Derealisation *T* | -0.02 | 0.89 | -0.31 | 0.03<sup>†</sup> | -0.08 | 0.60 |
| MDI Emotional Constriction *T* | 0.07 | 0.63 | -0.29 | 0.04<sup>†</sup> | -0.13 | 0.38 |
| MDI Memory Disturbance *T* | -0.10 | 0.51 | -0.59 | **<0.001** | -0.13 | 0.40 |
| MDI Identity Disturbance *T* | 0.36 | 0.01<sup>†</sup> | -0.36 | **0.01** | 0.14 | 0.36 |
| SF-36 Role Limitations – Physical | 0.01 | 0.94 | 0.54 | **<0.001** | -0.13 | 0.38 |
| **RVIP Ability** |  |  |  |  |  |  |
| IEDSS Total Errors | -0.24 | 0.15 | -0.48 | **0.002** | -0.16 | 0.30 |
| *IEDSS Total Trials* | -0.33 | 0.04<sup>†</sup> | -0.48 | **0.002** | -0.17 | 0.27 |
| *IEDSS Response Latency* | -0.23 | 0.16 | -0.35 | **0.03** | -0.32 | 0.04<sup>†</sup> |
| ***RVIP Probability of Hit*** |  |  |  |  |  |  |
| *IEDSS Total Errors* | -0.14 | 0.42 | -0.47 | **0.003** | -0.14 | 0.38 |
| *IEDSS Total Trials* | -0.24 | 0.15 | -0.42 | **0.008** | -0.15 | 0.33 |
| *IEDSS Response Latency* | -0.14 | 0.42 | -0.32 | **0.048** | -0.32 | 0.04<sup>†</sup> |
| ***RVIP Total Misses*** |  |  |  |  |  |  |
| *IEDSS Total Errors* | 0.14 | 0.42 | 0.47 | **0.003** | 0.14 | 0.38 |
| *IEDSS Total Trials* | 0.15 | 0.33 | 0.42 | **0.008** | 0.15 | 0.33 |
| *IEDSS Response Latency* | 0.14 | 0.42 | 0.32 | **0.048** | 0.32 | 0.04<sup>†</sup> |
| ***IEDSS Performance ratings*** |  |  |  |  |  |  |
| *IEDSS Total Errors* | -0.53 | **<0.001** | -0.38 | **0.02** | -0.09 | 0.58 |
| *IEDSS Total Trials* | -0.50 | **<0.001** | -0.40 | **0.01** | -0.10 | 0.52 |
| *IEDSS Response Latency* | -0.37 | **0.02** | -0.45 | **0.004** | -0.14 | 0.36 |

**Key:** CC=clinical controls; CFQ=Cognitive Failures Questionnaire; FMS=functional motor symptoms; FNSQ=Functional Neurological Symptoms Questionnaire; FS=functional seizures; GAD-7=Generalised Anxiety Disorder-7; IEDSS=Intra-Extra Dimensional Set Shift; MDI=Multiscale Dissociation Inventory; PHQ=Patient Health Questionnaire; RVIP=Rapid Visual Information Processing; SDQ-20=Somatoform Dissociation Questionnaire-20; SF-36=Short-Form Health Survey-36 item; WSAS=Work and Social Adjustment Scale <sup>†</sup>Did not survive Benjamini-Hochberg correction (5%)

